# Genome-wide Analysis of Rare Haplotypes Associated with Breast Cancer Risk: Discovery, Replication, and Generalizability Evaluation

**DOI:** 10.1101/2022.10.21.22281360

**Authors:** Fan Wang, Wonjong Moon, William Letsou, Yadav Sapkota, Zhaoming Wang, Cindy Im, Jessica L. Baedke, Leslie Robison, Yutaka Yasui

**Affiliations:** Department of Epidemiology and Cancer Control, St. Jude Children’s Research Hospital, Memphis, Tennessee 38105, USA; School of Public Health, University of Alberta, Edmonton, Alberta T6G 1C9, Canada

**Author notes:** **Corresponding authors** Fan Wang, Department of Epidemiology and Cancer Control, St. Jude Children’s Research Hospital, 262 Danny Thomas Place, Mail Stop 735, Memphis, TN 38105, USA. Yutaka Yasui, Department of Epidemiology and Cancer Control, St. Jude Children’s Research Hospital, 262 Danny Thomas Place, Mail Stop 735, Memphis, TN 38105, USA.

**Keywords:** Genome-wide association studies, Genetic interaction, Genetic heterogeneity, Rare variants

## Abstract

While numerous common variants have been linked to breast cancer (BCa) risk, they explain only partially the total BCa heritability. Inference from the Nordic population-based twin data indicates that rare high-risk loci are the chief determinant of BCa risk. Here, we use haplotypes, rather than single variants, to identify rare high-risk loci for BCa. With computationally phased genotypes from 181,034 white British women in the UK Biobank, we conducted a genome-wide haplotype-BCa association analysis using sliding windows of 5-500 consecutive array-genotyped variants. In the discovery stage, haplotype associations with BCa risk were evaluated retrospectively in the pre-study-enrollment portion of data including 5,487 BCa cases. BCa hazard ratios (HRs) for additive haplotypic effects were estimated using Cox regression. Our replication analysis included women free of BCa at enrollment, of whom 3,524 later developed BCa. This two-stage analysis detected 13 rare loci (frequency <1%), each associated with an appreciable BCa risk increase (discovery: HRs=2.84-6.10, P-value<5×10^−8^; replication: HRs=2.08-5.61, P-value<0.01). In contrast, the variants that formed these rare haplotypes individually exhibited much smaller effects. Functional annotation revealed extensive cis-regulatory DNA elements in BCa-related cells underlying the replicated rare haplotypes. Using phased, imputed genotypes from 30,064 cases and 25,282 controls in the DRIVE OncoArray case-control study, six of the 13 rare-loci associations proved generalizability (odds ratio estimates: 1.48-7.67, P-value<0.05). This study demonstrates the complementary advantage of utilizing rare haplotypes to capture novel risk loci and possible discoveries of more genetic elements contributing to BCa heritability once large, germline whole-genome sequencing data become available.

## Introduction

Common- and rare-variant models are two contrasting hypotheses under active debate regarding how germline genetic variants drive disease risk (1). In the past decade, efforts for identifying germline determinants of breast cancer (BCa) risk have been largely led by genome-wide association studies (GWAS) of single nucleotide polymorphisms (SNPs), which have identified hundreds of common risk variants in large samples (2,3). Polygenic risk scores comprised such GWAS-discovered risk variants have shown ability to grade women’s BCa risk by several fold (3,4). Inference from the Nordic twin data (5), however, clearly supports the rare-variant model over the common-variant counterpart, explaining the specific epidemiological patterns of BCa in the largest, population-based twin study of cancer (Yasui et al. submitted as a separate manuscript), consistent with McClellan and King’s hypothesis that genetic heterogeneity involving many rare mutations drives the risk of BCa and other common diseases (6).

GWAS methodologies for testing common-variant associations with disease risk are mature and have widely been applied. In GWAS, genotype imputation increases genomic coverage and allows researchers to combine multiple studies with potentially different genotyping platforms for higher statistical power (7). On the other hand, testing rare variant associations with disease risk across the genome in a sample of unrelated subjects has been challenging, if not infeasible, because doing so requires sample sizes large enough to identify rare-variant carriers and a genotyping platform appropriate for rare variants. Whole-genome sequencing (WGS) data from a large cohort would enable studies of rare variant associations. This important data collection is currently underway, but WGS data are still unavailable in large cohorts or case-control studies.

Here, we consider rare haplotypes, instead of rare, single variants, formed by sets of genotyped variants on an array in large epidemiological studies for identifying rare, genetic determinants of BCa risk. We submit two reasons for considering rare haplotypes: 1) multiple loci interact to exsert biological function (e.g., those of regulatory regions to control gene expression); and 2) each risk mutation must have arisen on a chromosome whose rare haplotypes may serve as proxies if unbroken by recombination over time (e.g., the mutation arose recently). Note, however, that this analysis does not consider haplotypes involving an ungenotyped variant and thus is far from a comprehensive examination of all rare haplotypes. Nonetheless, the novel rare haplotypes we identify provide further support for the rare-variant model and suggest consideration of haplotypes for the search for additional genetic contributions to BCa risk (and other common diseases).

Specifically, this paper reports genome-wide discovery and replication analyses involving haplotypes of variants genotyped in the UK Biobank (UKBB) (8) with its Axiom Array for BCa-risk association in 181,034 “white British” women, including 9,011 BCa cases. The UKBB findings were further evaluated using the Discovery, Biology, and Risk Inherited Variants in Breast Cancer (DRIVE) project (9) for generalizability evaluation with 30,064 cases and 25,282 controls of females with European genetic ancestry.

## Methods

### Study subjects

For discovery and replication analysis, we utilized the data from the UK Biobank (UKBB), a large cohort study that has collected extensive phenotypic and genetic data on approximately half a million participants across the United Kingdom (8). Upon accessing the UKBB resource under application number 44891, 181,034 women who self-reported as “white” and “British” with very similar genotype principal components (PCs) (the “white British ancestry subset” designated by UKBB) were included in our analysis. We excluded women who: 1) were identified to be an outlier in heterozygosity and missing genotyping rates; 2) showed any sex chromosome aneuploidies; 3) were related to other genotyped participants, as inferred by KING (30); or 4) withdrew from the UKBB before this study began. BCa incidence (UKBB Data-Field: 40006 and ICD-10 codes: C50.0 -C50.9) and other clinically relevant variables were extracted from the UKBB portal. Women selected for this study had ages between 40 and 71 years old at the time of enrollment, of whom 5,487 were diagnosed with BCa before enrollment at an average age of 52.9 years (standard deviation (SD)=7.4 years). This group was used in our discovery analysis. During follow-up after UKBB enrollment, 3,524 additional women developed BCa with an average age of 61.2 years (SD=7.7 years). This group was used in our replication analysis. The remaining 172,023 women without a BCa had an average age of 65.0 years (SD=7.9 years) at the last follow-up with a median follow-up time of 8.5 years.

To evaluate the generalizability of our UKBB findings (“generalizability analysis”), we used subjects in the DRIVE study, which started in 2010 as part of the GAME-ON initiative (https://epi.grants.cancer.gov/gameon/) to detect new genetic loci for five common cancer types using the Illumina custom OncoArray (9). The DRIVE study comprised 17 studies and collected 60,015 participants from 15 countries. We accessed the DRIVE OncoArray dataset through the NIH dbGaP (Study Accession: phs001265.v1.p1) and removed 4,669 (7.8%) participants based on our genotype quality control (QC) and limited the sample to those of European genetic ancestry (see below for details). A final set of 55,346 women, including 30,064 BCa cases and 25,282 controls, was assembled as an independent, slightly broader genetic-ancestry population than UKBB’s “white British” for assessing the generalizability of our findings from the UKBB discovery and replication analyses. All women had ages between 18 and 91 years old at the recruitment, and the BCa case group had an average age of onset of 61.7 years (SD=10.7 years). For both UKBB and DRIVE, all participants provided written informed consent under local IRB-approved protocols.

### The UKBB Phased Genotype Data

The current study utilized the phased genotype data provided by the UKBB as the output of whole-chromosome phasing during the genotype imputation process. Details regarding the UKBB’s Axiom Array, haplotype estimation, and genotype imputation have been previously described by UKBB scientists (8). Briefly, the phased data downloaded from UKBB included 487,409 participants phased on a per-chromosome basis at 658,720 autosomal genotyped variants with SHAPEIT3 (31) using the 1000Genome Phase 3 haplotypes (32) as the reference. We converted the supplied phased data in the BGEN v1.2 format to a new data structure, called GDS (33), to facilitate genome-wide haplotype analysis. Then we created a subset of the phased data for the 181,034 women selected for our analysis. The UKBB phased data had a 100% genotyping rate on all variant sites; therefore, the only exclusions we made were 12,274 variants that were either monomorphic (i.e., minor allele frequency (MAF) = 0) or failed the Hardy-Weinberg equilibrium (HWE) test (i.e., *P* <1×10^−10^) in our analytic subset. After QC, 646,446 variants (643,307 SNPs and 3,139 insertion/deletions) remained for analysis. The above data processing was implemented with PLINK (34) and R package “SeqArray” (33).

### The DRIVE OncoArray Data and Genotype Imputation

We obtained the DRIVE OncoArray data, containing 528,620 variants genotyped in 60,015 BCa cases and controls, from dbGaP under our approved project #28544. Given the limited overlap of variants between the UKBB’s Axiom Array and the DRIVE’s OncoArray, we first filtered the DRIVE genotype data through variant- and sample-QCs, as described below, and then performed genotype imputation to improve DRIVE coverage of the UKBB’s genotyped variants. We filtered out variants based on the following filters: 1) sex chromosome aneuploidies; 2) missing genotyping rate >10%; 3) MAF=0; and 4) HWE test *P*< 1×10^−10^. We excluded samples who: 1) had a genotyping rate <90%; 2) failed in the PLINK’s sex check (i.e., F-score above 0.3); 3) were related to any other samples with second-degree or closer relationship, as detected by KING (30); or 4) separated from European-ancestry samples in the 1000 Genomes Phase 3 data (32), according to the first two PCs computed by flashPCA (35). Women identified as of European ancestry in this study had PC1 ≤0.0025 and PC2 ≥-0.007, as indicated by vertical and horizontal dotted lines in **Suppl. Fig. S3B**. The cleaned data contained 433,297 variants and 55,450 unrelated women of European genetic ancestry. Besides, we performed the second PCA on the 55,450 women and the derived top 10 PCs were used to control the fine-scale population structure.

Genome-wide imputation of the cleaned DRIVE genotype data was carried out using the online Trans-Omics for Precision Medicine (TOPMed) Imputation Server (https://imputation.biodatacatalyst.nhlbi.nih.gov), which automatically executes its own QC, phasing with Eagle2 (36), and genotype imputation with Minimac4 (37) on a per-chromosome basis. The TOPMed pipeline was built on a large reference panel of 194,512 haplotypes and 308 million variants generated by the TOPMed program of the US National Heart, Lung and Blood Institute (38). Due to the sample limit of the server, the QCed DRIVE OncoArray data were split randomly into three subsets. Each subset contained 15,000-20,000 samples and was submitted for genome-wide imputation. After obtaining three sets of imputed data, we calculated the Rsq quality metric for each variant among the 55,450 women according to the Minimac formula (37): 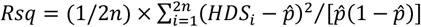 where 2*n* denotes the total number of phased chromosomes with the variant (among n women), *HDS* indicates the estimated haploid alternate allele dosage (between 0 and 1) for the variant on the *i-*th chromosome, and 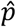 is the observed alternate allele frequency. Rsq ranges from 0 to 1 with values close to 1 indicating high imputation quality. Variants with Rsq≥0.8 were used for our DRIVE analysis. Moreover, 104 women of unknown age (code=888) were removed. In the end, we generated a subset of 615,181 phased imputed variants on 55,346 women, containing 95% of UKBB-phased variants, for the generalizability analysis.

### Genome-wide Haplotype Analysis: Discovery and Replication

Genome-wide haplotype analysis for BCa risk was carried out using the UKBB-phased genotypes under the following discovery-replication framework. In the discovery stage, haplotypes were directly constructed from a set of consecutive genotyped variants in windows of fixed size W = 5, 10, 20, 30, 50, 100, 250, and 500 variants. For each chromosome, we shifted the window by one variant in the 5’ -> 3’ direction. As such, our analysis was designed to cover every set of 5-500 consecutive genotyped variants. Each haplotype was coded as 0, 1, or 2 copies per person in the same way as SNPs, and its additive effect was tested statistically. The number of haplotypes observed in a window varied depending on the window size and the underlying linkage disequilibrium (LD) pattern. In the discovery analysis, each haplotype was tested for association with BCa risk in the pre-enrollment portion of the UKBB data, consisting of 5,487 BCa cases and 175,547 women free of BCa at study enrollment. To reduce the unnecessary computational burden, our discovery analysis tested only the haplotypes with frequency ≥ 0.1% (10 per 10,000 chromosomes) in the 5,487 pre-enrollment cases. For each haplotype, the hazard ratio (HR) of developing BCa was estimated by multivariable Cox regression with age as the time axis and right censoring at the study enrollment, controlling for the first ten genotype PCs. The analysis was implemented using R packages “survival” (https://CRAN.R-project.org/package=survival) and “SeqArray” (33). Statistical significance for the discovery analysis was set at Wald-test p-values of Cox regression (Cox *P*)<5×10^−8^. The genome-wide scan was performed in parallel on 22 autosomes with chunks of 500 windows for each fixed size. All runs were completed on our institutional high-performance computing cluster with 20 CPU cores per chromosome and 4Gb memory per core.

In the replication analysis, haplotypes that met the statistical significance threshold of the discovery analysis were evaluated in the post-enrollment part of the UKBB data, excluding the 5,487 pre-enrollment BCa cases. This replication analysis included 3,524 post-enrollment-onset BCa cases and 172,023 women without a BCa at the last UKBB follow-up before our UKBB data access. Haplotypes’ BCa HRs were estimated by the same multivariable Cox model, taking age as the time axis starting from the study enrollment to the last follow-up and controlling for the first ten genotype PCs. The replication statistical significance threshold was set at Cox *P*<0.01: see below for an assessment of false discovery/replication by the statistical significance thresholds we used.

### Lead Haplotype among Correlated Haplotypes

Because of our use of overlapping windows, many discovered/replicated haplotypes are highly correlated/redundant. To consolidate each set of these correlated haplotypes into a single risk locus (or region) represented by a “lead haplotype”, we merged replicated haplotypes by LD-based clumping using PLINK (34) with an LD threshold of *r*^2^>0.1 and a physical distance threshold of 500 kb. The position of the first variant on each replicated haplotype was used as the haplotype’s genomic position. Independent risk loci/region were determined as the mutually exclusive clumps formed by the clumping procedure. We calculated the overall HR and Cox likelihood ratio test (LRT) p-value for each replicated haplotype, using the entire set of UKBB data (combining the discovery and replication data into a total of 9,011 BCa cases and 172,023 BCa-free women). Within each risk locus/region, the haplotype exhibiting the smallest p-value was considered the “lead haplotype”.

### Exploration of False Discovery Rate by Permutation Experiment

Because we used highly overlapping windows and tested many correlated/redundant haplotypes, it is not clear whether our statistical significance thresholds, *P*<5×10^−8^ for discovery and *P*<0.01 for replication, adequately controlled the false discovery rate. To assess the extent of potential false discovery in the discovery-replication framework with the UKBB data, we conducted a permutation experiment. Three permuted datasets were generated in which the phenotype vectors (i.e., the vectors of BCa diagnosis status, age variables, and genotype PCs) were randomly shuffled among the 181,034 women and used with their original unshuffled genetic data vectors. This simulated the null hypothesis condition where there is no association between BCa and haplotypes, except by chance, while keeping the original associations of age and the PCs with BCa. For each permuted dataset, we repeated the same two-stage haplotype analysis we conducted above, except for W=500 variants (there was no replicated haplotype of W=500 in the original analysis). The number of haplotypes that met the same statistical significance thresholds (*P*<5×10^−8^ for discovery and *P*<0.01 for replication) and the number of independent risk loci/region after the LD-based clumping were used as approximate estimates of the numbers of false discoveries we might expect under the null hypothesis of no association.

### Individual Variant Association with BCa risk

In addition to the haplotype analysis, we performed a standard GWAS analysis to assess individual variants’ associations with BCa risk using the UKBB data. For each variant, the BCa-genotype association was assessed under the additive model using multivariable Cox regression, in the UKBB data of 9,011 BCa cases and 172,023 BCa-free women. Age was taken as the time axis and the model controlled for the first ten genotype PCs. The GWAS was completed parallelly on 22 autosomes in chunks of 200 variants using 10 CPU cores and 4Gb memory per core.

### Generalizability Analysis

In assessing the generalizability of the replicated findings, a large, independent BCa dataset with phased genotype data that contained the UKBB genotyped variants would have been ideal. In the absence of such a dataset, we used the cleaned DRIVE BCa case-control data for generalizability evaluation with statistical phasing and imputation performed as described above. Specifically, we assessed the lead haplotype of each risk locus/region identified by our UKBB two-stage discovery/replication analysis in the QCed DRIVE imputed data including 30,064 BCa cases and 25,282 BCa-free controls.

Because statistical phasing and imputation of the DRIVE data is subject to errors, the following strategy was used. First, we utilized pairs of “HDS” values (“estimated phased haploid alternative allele dosages”) available for each variant of each woman in the final phased/imputed DRIVE data. The two HDS values for a variant of a woman denote the two probabilities of the alternative allele on her two phased haplotypes. The alternative allele was called when the HDS value was above 0.5; otherwise, the reference allele was called. Second, we reduced the number of variants required to define each lead haplotype to avoid the impact of potential imputation errors in unnecessary variants. Specifically, for the *i-*th replicated lead haplotype defined by W_i_ consecutive UKBB variants, we included the subset of W_i_ variants that passed the imputation-accuracy filter (Rsq≥0.80) and applied tree-based recursive partitioning, implemented by R package “rpart”, to reduce the number of variants necessary to define the lead haplotype to m_i_≤Wi variants, with the complexity parameter (CP) 10^−10^ and the minsplit parameter 40 (and the other parameters at their default values). At one locus/region (Locus #19) where the reduced variant set did not approximate the lead haplotype well, we relaxed the Minimac filter to Rsq≥0.3 for common variants with a MAF ≥ 1% and reran the variant reduction for the new lead haplotypes.

The evaluation scheme above (hereafter referred to as “original haplotype calls”) was our primary scheme for defining each DRIVE woman’s diplotypes of lead haplotype i. which can be expressed as:

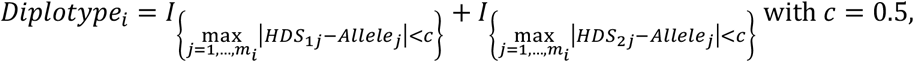

where “Allele” is the code (0 or 1) for the allele of the lead haplotype, and the first index “1” or “2” of HDS denotes the woman’s first or second chromosomes. Note that |*HDS*_1*j*_ − *Allele*_*j*_| and |*HDS*_2*j*_ − *Allele*_*j*_| indicate imputation uncertainty of the *j*-th variant of the reduced variant set of the *i*-th lead haplotype, with possible values between 0 and 1. Values close to 0 and 1 indicate high confidence in the agreement or disagreement, respectively, of the imputed allele with the allele in the lead haplotype, with the boundary 0.5 corresponding to an ambiguous imputation result.

To account for the imputation uncertainty in the generalizability analysis more conservatively, we supplement our primary scheme with the following sub-schemes for defining diplotypes in DRIVE. The secondary scheme, “high-confidence haplotype calls,” considered only variants imputed with high confidence by using a smaller value of *c* (considering *c* in the range 0–0.01 with an increment of 0.001 and 0.01–0.50 with an increment of 0.01) and reporting the most statistically significant association. The tertiary scheme, “high-confidence haplotype calls with one exception,” removed one variant from the reduced variant set of the lead haplotype and applied the secondary scheme to the remaining variants in the set (with the excluded variant still satisfying |*HDS* − *Allele*|< 1).

The primary, secondary, and tertiary schemes were applied sequentially. The association between each imputed lead haplotype and BCa case/control status in DRIVE was assessed by logistic regression, adjusting for age and the first ten genotype PCs, where likelihood-ratio test (LRT) *P*<0.05 served as the statistical significance measure of the association. Fisher’s Exact Test was also conducted to supplement the LRT as the frequencies of lead haplotypes were low.

### Functional Annotation

To explore the biological functions of each BCa-risk haplotype, we annotated variants on the rare lead haplotypes that met all three statistical significance thresholds of the discovery, replication, and generalizability evaluation, using a variety of functional datasets (see Data Availability below for URLs). UCSC LiftOver chain files were used to convert genomic coordinates between hg19 (i.e., DRIVE OncoArray data) and hg38 (i.e., DRIVE TOPMed-imputed data). Gene-based annotations, such as nearest genes and functional consequences, were extracted using WGSA v0.85 (39) with GENECODE v37. Regulatory variants were annotated by searching the following resources: 1) *cis*-eQTLs reported in GTEx v8 database, passing 5% false discovery rate in the breast mammary tissue (16); 2) Human active enhancers in the HACER (18) and enhanceratlas2 (17), limiting to five BCa-related cell/tissues (i.e., MCF-7, MCF10A, HCC1806, HMEC, and T47D); 3) Enhancers experientially characterized by a STARR-seq in the MCF-7 cell line (ENCODE accession: ENCSR547SBZ), where significant enhancer peaks were defined by *P*<0.05; 4) Topologically Associating Domains (TADs) and chromatin loops predicted by the 3D Genome browser, where variants with putative regulatory roles were considered within 20Kb of a TAD boundary (20) or a chromatin-loop anchor regions where DNA segments physically interact (21); and 5) DNA-binding motifs from HaploReg v4 (19), where we considered variants to be regulatory if the absolute change of the reported LOD score between reference and alternative alleles (|LOD_alt_-LOD_ref_|) was above 3, with a positive value of LOD change indicating that the alternate allele likely resulted in stronger motif binding and a negative change indicating that the alternative allele likely caused weaker motif binding. We combined the above annotations to summarize potential functional features at the haplotype level.

### Data availability statement

The data generated in this study are available within the article and its supplemental data files. Individual-level genotype and phenotype data are publicly available by submitting request to the UK Biobank and dbGaP (study accession: phs001265.v1.p1). Analytic scripts and codes are available at https://github.com/fwplace/HapProj.

Public databases and software used in this study include:

LDlink: https://ldlink.nci.nih.gov/?tab=home

GWAS catalog: https://www.ebi.ac.uk/gwas/

TOPMed Imputation server: https://imputation.biodatacatalyst.nhlbi.nih.gov

UCSC chain files: https://hgdownload.cse.ucsc.edu/goldenpath/hg19/liftOver/

GTEx eQTL v8: https://gtexportal.org/home/

Recombination rate: https://github.com/cbherer/Bherer_etal_SexualDimorphismRecombination

GENECODE v37: https://www.gencodegenes.org/human/release_37lift37.html

STAR-seq in the MCF-7 cell line: https://www.encodeproject.org/

HACER: http://bioinfo.vanderbilt.edu/AE/HACER/index.html

Enhanceratlas2: http://www.enhanceratlas.org

3D Genome Browser: http://3dgenome.fsm.northwestern.edu/publications.html

HaploReg v4.1: https://pubs.broadinstitute.org/mammals/haploreg/haploreg.php

R survival package: https://CRAN.R-project.org/package=survival

R SeqArray package: https://bioconductor.org/packages/release/bioc/html/SeqArray.html

## Results

### Genome-wide Haplotype Analysis Using the UKBB Phased Data

The overall design of our genome-wide haplotype analysis is shown in **Fig. 1A**. The discovery analysis included 181,034 female participants who were classified as ‘white British’ by the UKBB (see Methods for inclusion/exclusion criteria). All women were biologically unrelated and had a homogeneous genetic background consistent as a specific subgroup of European genetic ancestry in comparison to the 1000 Genomes Phase 3 European population samples (**Suppl. Figure S1**). The set of phased haplotypes from UKBB for the 181,034 women contained 646,446 genotyped variants, enabling the construction of haplotypes genome-wide through sliding windows of 5, 10, 20, 30, 50, 100, 250, and 500 consecutive genotyped variants. For each window size, 4.8–88.4 million haplotypes that had frequency ≥ 0.1% (i.e., 10 per 10,000 chromosomes) in 5,487 pre-enrollment BCa cases were identified and individually tested for BCa-risk associations by Cox regression with age as the time axis and right censoring at study enrollment, adjusting for the first ten genotype PCs. The genome-wide scans detected 5,858 haplotypes across the eight fixed-size sliding windows at *P*<5×10^−8^ (**Table 1**).

**Figure 1.**
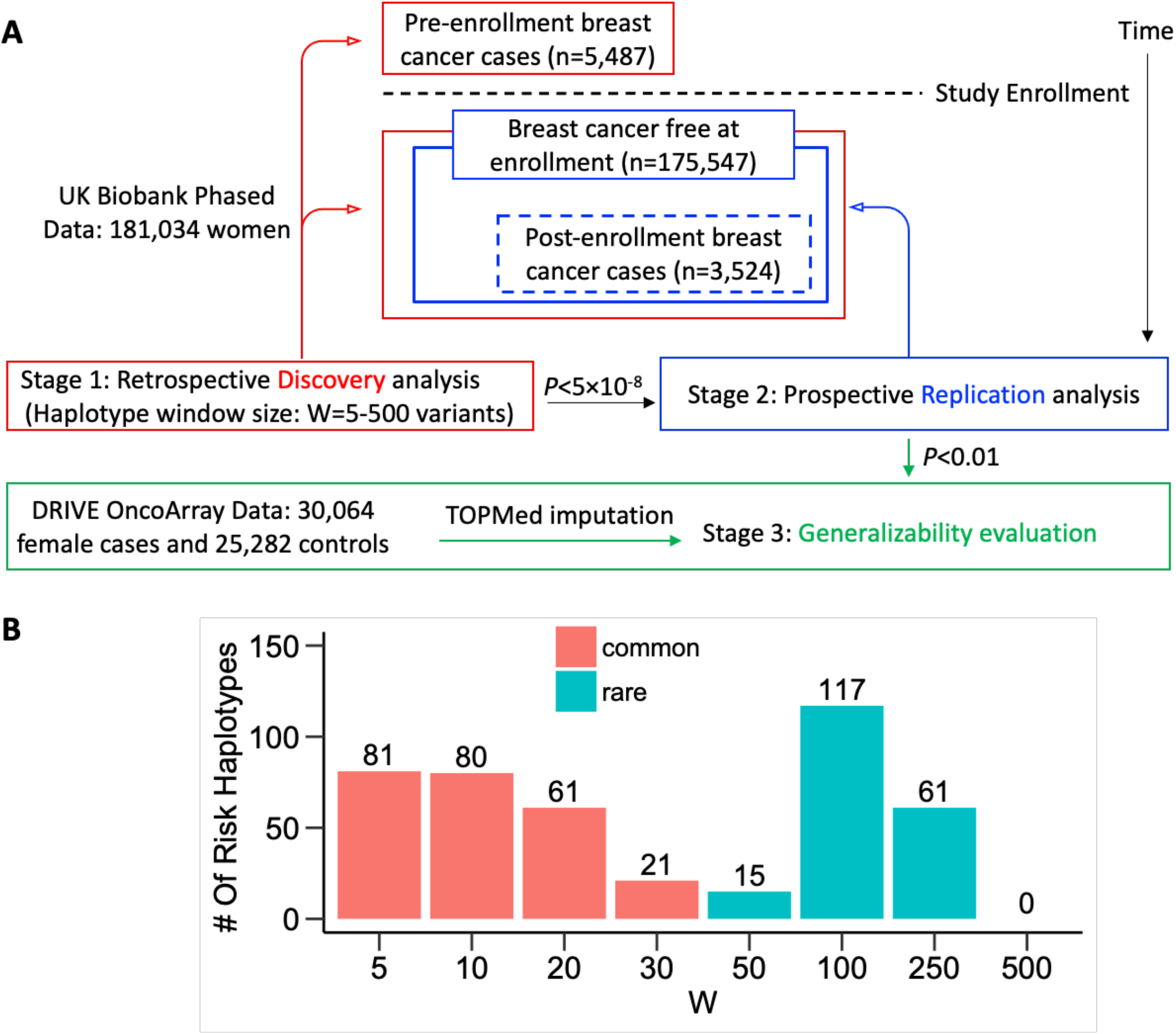
The Overall Design of Genome-wide Haplotype Analysis for BCa. (A) Flowchart of the two-stage haplotype analysis using the UK Biobank dataset and generalizability evaluation using the DRIVE dataset. (B) Counts of common (frequency ≥ 1%) and rare (frequency < 1%) that met the discovery (*P*<5×10^−8^) and replication (*P*<0.01) statistical significance threshold in the UK Biobank phased data under various window sizes of W=5–500 consecutive genotyped variants.

**Table 1.** Summary of the Two-stage Haplotype Analysis Results Using the UK Biobank Phased Data.

| Window Size <sup>a</sup> | Effective Windows | Genomic Length | Discovery Analysis |  |  | Replication Analysis |  |
| --- | --- | --- | --- | --- | --- | --- | --- |
| | | | Coverage, % <sup>b</sup> | Total Tests <sup>c</sup> | $P < 5 \times 10^{-8}$ | $P < 0.01$ | No. of Loci <sup>d</sup> |
| W=5 | 646,328 | 17.2 Kb | 41.272 | 4,802,633 | 90 | 81 | 7 |
| W=10 | 646,248 | 38.8 Kb | 20.420 | 12,237,248 | 117 | 80 | 6 |
| W=20 | 646,028 | 81.8 Kb | 8.350 | 34,466,021 | 215 | 61 | 5 |
| W=30 | 645,808 | 124.9 Kb | 4.338 | 58,110,973 | 473 | 21 | 3 |
| W=50 | 645,368 | 211.0 Kb | 1.611 | 88,427,869 | 995 | 15 | 5 |
| W=100 | 644,263 | 426.4 Kb | 0.338 | 85,988,703 | 1,769 | 117 | 9 |
| W=250 | 618,760 | 1.08 Mb | 0.034 | 25,950,002 | 1,673 | 61 | 3 |
| W=500 | 283,807 | 2.52 Mb | 0.007 | 3,426,133 | 526 | 0 | 0 |

<sup>a</sup> W, the number of consecutive genotyped variants that define the haplotypes. Each haplotype window is shifted by one variant.
<sup>b</sup> Coverage indicates the fraction of total haplotypes constructed in sliding windows that were tested in this study
<sup>c</sup> The number of haplotypes meeting the criterion of frequency $\geq 0.1\%$ among 5,487 pre-enrollment BCa cases.
<sup>d</sup> Independent loci/regions were determined by PLINK LD clumping with $r^2 > 0.10$ and a maximum distance of 500 kb between the lead haplotype and the other correlated haplotypes.

The 5,858 haplotypes from the discovery analysis of the retrospective component of the UKBB data were evaluated in the following replication analysis among the BCa-free women at their UKBB enrollment (i.e., the non-cases in the discovery analysis), following them prospectively for BCa incidence starting from the enrollment. Note that, while the replication analysis used the same UKBB cohort, it is independent of the discovery analysis because all women in the replication analysis were non-cases in the discovery analysis, were BCa free at the start of the prospective replication analysis, and the replication analysis utilized case/non-case data that emerged after the UKBB study enrollment, i.e., after the completion of case/non-case data utilized in the retrospective discovery analysis. Using the same Cox regression with 3,524 post-enrollment BCa cases among 175,547 women BCa-free at the enrollment, 436 haplotypes replicated at *P*<0.01 (**Table 1**). These replicated haplotypes were split into two mutually exclusive groups: one containing 243 common short haplotypes with a window size of 5-30 variants, and the other containing 193 rare long haplotypes with a window size of 50-250 variants (**Fig. 1B**). For windows of 500 variants, no haplotype replicated.

To explore the false discovery rate of our discovery-replication analyses of haplotypes with the UKBB phased data, we ran three permutation experiments by randomly permuting the BCa status of 181,034 women together with age and genotype PCs, while keeping the original genetic data, to create the null hypothesis condition of no BCa-haplotype association. Then we performed the identical genome-wide discovery and subsequent replication analyses of the permuted dataset with the same statistical analysis and significance thresholds. While the permuted discovery analysis yielded similar numbers of genome-wide significant haplotypes as the original discovery analysis, indicating that many of the “original-analysis discoveries” are likely false positives (**Suppl. Table S1**), the subsequent replication analysis showed very few replications, indicating the replicated haplotypes of the original analysis are mostly valid (**Suppl. Table S1**).

### Prioritization of 13 Rare Haplotype Loci for BCa Risk

To remove redundancies in the replicated haplotypes, we applied LD clumping to the 436 replicated haplotypes to consolidate each set of correlated/overlapping haplotypes into a single risk locus/region. After LD clumping, the replicated haplotypes were grouped into 20 distinct genetic loci/regions, each being represented by a “lead haplotype” whose BCa-association p-value, computed by the combined Cox-regression analysis of 9,011 (pre-enrollment + post-enrollment) BCa cases and 172,023 BCa-free controls, was the smallest among the replicated haplotypes of the same locus/region (**Suppl. Table S2**). Of the 20 risk loci/regions, seven (Locus IDs: # 1-4, 7, 8, and 10) were mapped by common haplotypes with a window size of 5 variants and genomic length of 5.2-42.2 kb. The seven common lead haplotypes (i.e., those of *FGFR2* on 10q26.3, *CASC16* on 16q12.1, *DIRC3-AS1* on 2q35, *LINC01488/CCND1* on 11q13.3, *C5orf67/MAP3K1* on 5q11.2, *NEK10* on 3p24.1, and *CASC8/CASC21* on 8q24.21) conferred a modest BCa risk among 181,034 women (hazard ratio (HR)=1.11-1.30, *P*=2.3×10^−67^-4.3×10^−12^, **Suppl. Table S3**). These common haplotypes appeared to be tagged by their respective lead variants of standard GWAS analyses (HR=1.11-1.30, *P*=4.1×10^−66^-2.6×10^−5^) or be in tight LD with known GWAS hits (**Suppl. Table S3**).

In contrast, the remaining 13 loci (Locus IDs: # 5, 6, 9, and 11-20) were mapped by rare haplotypes with larger window sizes covering longer genomic regions. As shown in **Table 2**, the 13 rare lead haplotypes exhibited relatively large BCa risk with HRs of 2.84-6.10 in the discovery analysis and HRs of 2.08-5.61 in the replication analysis. From the standard GWAS analysis, no variants within ± 400 kb of the 13 rare haplotypes were found to pass the genome-wide significance threshold (i.e., *P*<5×10^−8^) for association with BCa risk, with the lowest p-values ranging between 4.1×10^−4^ and 0.040. The rare haplotype on 22q12.1 (Locus #5), formed by 250 consecutive genotyped variants spanning in a region of 1.4 Mb, demonstrated the most statistically significant association with BCa risk (frequency=0.13%, HR=3.49, *P*=1.5×10^−15^, **Fig. 2A**). The region contained multiple, known GWAS hits linked to the *MN1/PITPNB/TTC28/CHEK2/KREMEN1* gene cluster (3). However, only weak BCa-risk associations were observed with individual variants among the 181,034 UKBB women: the strongest association was rs35313550 in the intron 2 of *TTC28* (minor allele frequency (MAF)=2.5%, HR=0.85, *P*=4.1×10^−4^). Similarly, four additional rare haplotypes (Locus IDs: #14-16, and 18) spanned a region with known GWAS-hits/genes (3,10,11): *GRIN2A/ATF7IP2* on 16p13.2 (50 variants, length=106 kb, HR=2.93, *P*=2.2×10^−10^), *FRY/BRCA2* on 13q13.1 (100 variants, length=361 kb, HR=4.11, *P*=2.8×10^−10^), *AC107463*.*1/LINC02485* on 4q28.3 (100 variants, length=656 kb, HR=4.42, *P*=9.4×10^−10^), and *KIAA1217* on 10p12.2 (100 variants, length=523 kb, HR=4.44, *P*=2.6×10^−10^) (**Fig. 2 G-I, K**). To the best of our knowledge, the remaining seven rare haplotype loci (Locus IDs: #6, #9, #11-13, #17, #19, and #20) have not been identified for BCa risk: they are *C1QTNF3-AMACR/RAI14* on 5p13.2 (100 variants, length=658 kb, HR=5.91, *P*=5.0×10^−14^); *LSAMP* on 3q13.31 (100 variants, length=428 kb, HR=4.54, *P*=4.4×10^−11^); *FANC1/POLG* on 15q26.1 (250 variants, length=772 kb, HR=5.18, *P*=4.9×10^−11^); *GOT2/SLC38A7* on 16q21 (100 variants, length=519 kb, HR=4.75, *P*=1.5×10^−10^); *LINC02141/AC00981*.*2* on 16q21 (100 variants, length=487Kb. HR=4.70, *P*=2.0×10^−10^); *AC073062*.*1/AC016730*.*1* on 2p24.3 (50 variants, length=211 kb, HR=2.53, *P*=2.1×10^−9^); *MYB/AHI1* on 6q23.3 (100 variants, length=493Kb, HR=3.68, *P*=7.5×10^−11^); and *PARD3* on 10p11.21 (250 variants, length=1.1 Mb, HR=3.62, *P*=3.1×10^−11^) (**Fig. 2B-F, J, L-M**).

**Table 2.**
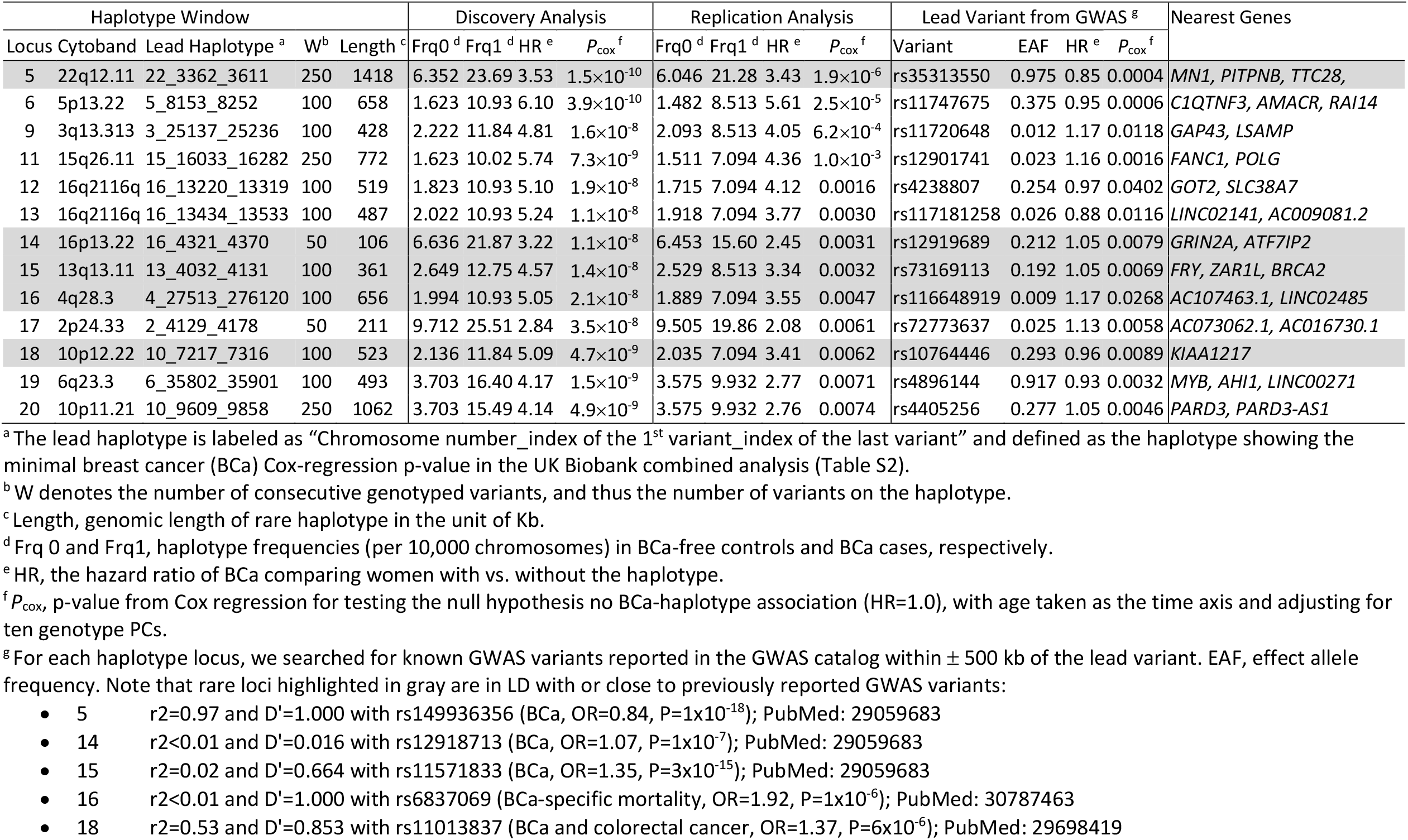
Identification of Thirteen Rare Haplotype Loci for Breast Cancer Risk in the UK Biobank Phased Data.

**Figure 2.**
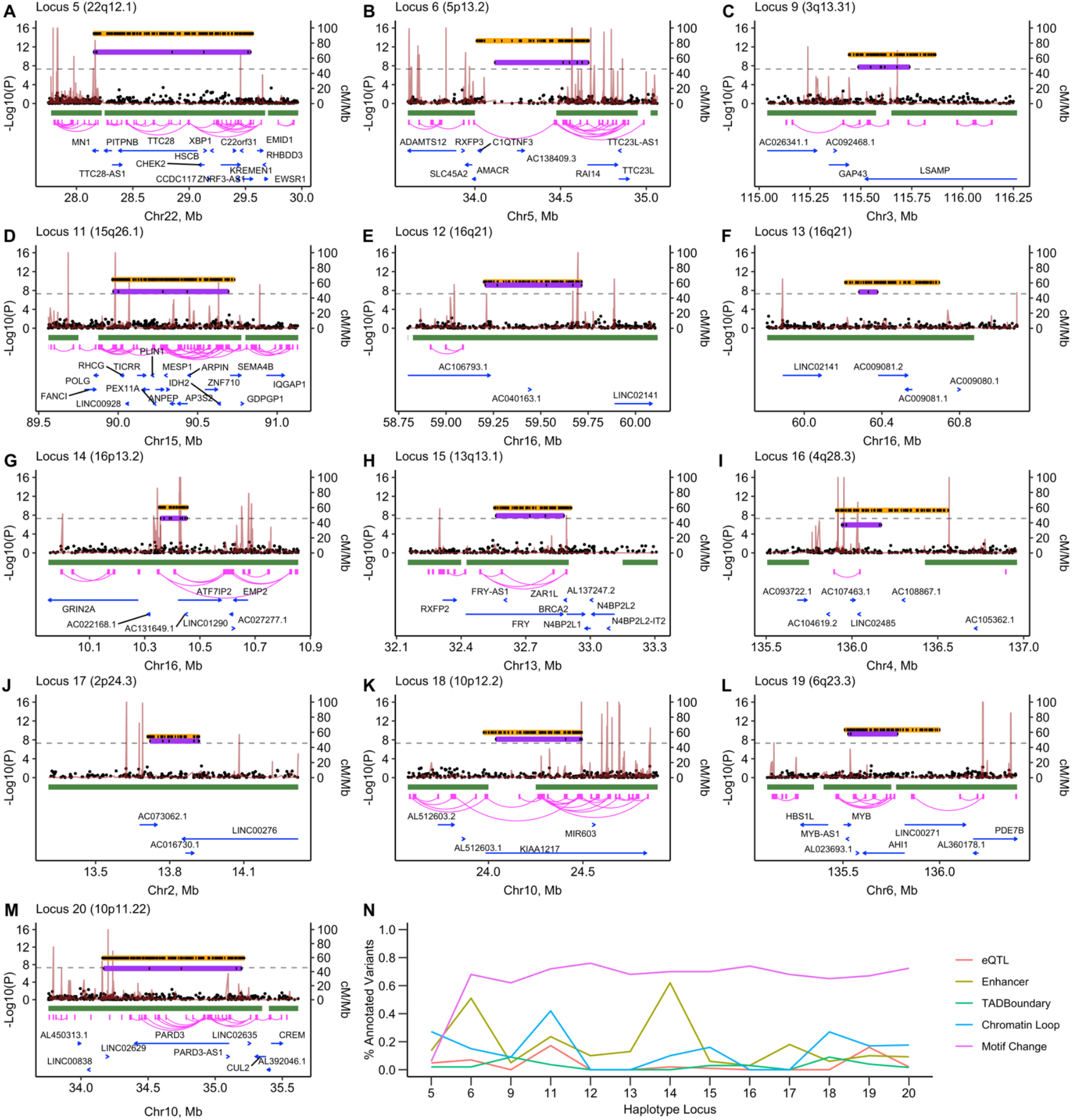
Genetic Association Plots of 13 BCa Rare Haplotypes Identified in the UK Biobank data. (A-M) Plots of the 13 loci with rare haplotypes that met the discovery (*P*<5×10^−8^) and replication (*P*<0.01) statistical significance thresholds in the UK Biobank phased data (also listed in Table 1). Each panel is labeled by its lead haplotype (“Chromosome number_index of the 1^st^ Variant_index of the last Variant”) and cytogenetic location. The left y-axis indicates association p-values for the original haplotype (orange segment) the reduced haplotype (purple segment), and individual variants from the standard GWAS analysis (black dots), calculated by Cox regression using all “white British” UK Biobank women. The genomic coordinates shown on the x-axis are based on GRCh37. The brown line shows the recombination rate computed by Bhérer *et al*. 2017 among European females, as indicated on the right y-axis. The bottom annotation tracks show locations of TADs (green segments), chromatin loops (magenta curves), and genes collected from 3D Genomes Browser and GENECODE v37. (N) the fraction of functionally annotated variants on rare haplotypes (see the Method Section for details).

To characterize the 13 rare loci, we analyzed the underlying LD-block patterns using a graphical-model-based LD partition method Big-LD (12). The Big-LD detected LD blocks (formed by a subset of highly correlated variants) and orphan variants (those not correlated with the other variants in the region) for each of the 13 rare loci (**Fig. 3**). The frequencies of haplotype segments formed by variants in single LD blocks are often higher than those of the original rare haplotypes, indicating that the rare lead haplotype may have arisen from multiple LD blocks via recombination, except for Locus #13 which might have arisen from a single small LD block (**Fig. 3F**). We tried to identify critical regions of each rare lead haplotype by excluding 20% of the consecutive variants and evaluating BCa-risk associations with truncated haplotypes that were formed by the remaining variants. We found that exclusion of variants on 5’ and 3’ ends led to a reduction of statistical significance of the identified rare haplotypes’ associations with BCa risk (**Suppl. Fig. S3 and Table S4**). Moreover, functional annotation with public databases revealed that a large proportion of variants on the rare lead haplotypes exhibited putative roles in regulating gene expression, enhancer activity, or 3D chromatin interactions (**Suppl. Table S5**). Of note, many annotated variants resided in the anchor regions of human chromatin loops and were predicted to alter bindings of DNA motifs (**Fig. 2N**).

**Figure 3.**
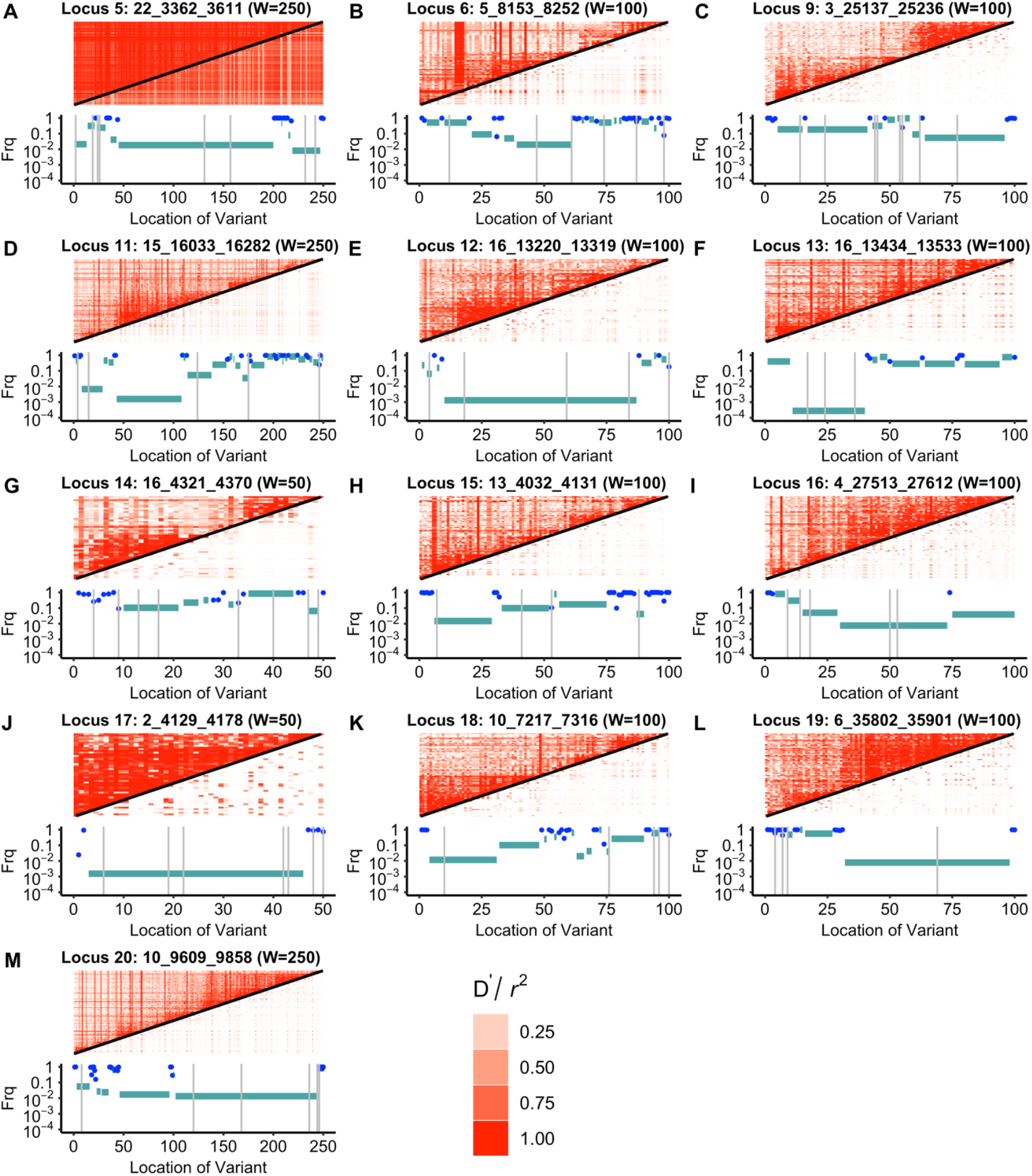
LD Heatmap Plots of 13 Rare Haplotypes Identified in the UK Biobank Data. Panels A-M correspond to the 13 lead risk haplotypes in **Table 2**. In each panel, the upper heatmap plot displays pair-wise D’ (upper triangle) and *r*^2^ (lower triangle) values computed using phased genotypes of 181,034 women. The heatmap color key is shown in the last row. The lower plot depicts the estimated LD structure calculated by the Big-LD algorithm (Kim et al. 2018) with |*r*| ≥ 0.50 and a maximal distance of 1 Mb between any two correlated variants. The y-axis shows the frequencies of truncated rare haplotypes (whose alleles match those of the original haplotype; cyan segments) and orphan variants (which were not in LD with other variants; blue dots). The x-axis represents the SNP index, and the vertical gray lines indicate the reduced variant sets used in the generalizability evaluation.

### Generalizability Evaluation of 13 Rare Lead Haplotypes for BCa Risk Using the DRIVE OncoArray Data

We sought to assess the generalizability of the associations of 13 rare lead haplotypes with BCa risk in an independent sample of 30,064 BCa cases and 25,282 BCa-free controls from the DRIVE study. All 55,346 women included in this analysis were of European genetic ancestry (i.e., those had PC1 ≤0.0025 and PC2 ≥-0.007 and were separated by dotted lines in **Suppl. Fig. S3B**), for which the “white British” of the UKBB cohort is a homogeneous subgroup (**Suppl. Fig. S1**) and thus we refer this analysis “generalizability evaluation” to a broader group. DRIVE samples were genotyped using the Illumina OncoArray. Because only 12% of genotyped variants in the UKBB phased data were included in the OncoArray, we performed a genome-wide imputation of the DRIVE OncoArray genotypes to improve coverage using the TOPMed Imputation Server (see Methods for more details). After post-imputation QC, we obtained high-quality phased, imputed genotypes on 1,304 (i.e., Minimac Rsq ≥0.80) of the 1,650 UKBB variants that formed the 13 rare lead haplotypes (**Suppl. Fig. S4A**). While MAFs of these individual variants were highly consistent between UKBB and DRIVE (**Suppl. Fig. S4B**), our analysis of haplotypes was subject to potential phasing/imputing errors. To alleviate this problem, we took the following strategy.

First, using the UKBB data, we reduced the set of variants within the window of each rare lead haplotype to a minimum subset required to define the lead haplotype by limiting to well-imputed variants (i.e., Minimac Rsq ≥0.80 in DRIVE) only and applying recursive partitioning, implemented in the R package “rpart”, so that phasing/imputation errors of unnecessary variants would not affect the analysis. As shown in **Fig. 2**, 13 rare lead haplotypes were well approximated by their minimum subsets consisting of 3-17 variants, much smaller numbers of variants than what their original haplotype windows contained. We re-evaluated the associations of the reduced rare lead haplotypes with BCa risk in the UKBB data and confirmed little change in their associations from the original lead haplotype (i.e., p-values became slightly less significant due to the approximation) (**Fig. 2A-M**). The exception was locus #19 where the lead haplotype could not be approximated. For this locus, we relaxed the Rsq filter to ≥0.30 for common variants (i.e., MAF ≥1%): this resulted in a reduced rare lead haplotype of 4 variants which yielded a similar association as the original lead haplotype (**Fig. 2L**). With LD analysis, we found consistently small *r*^2^ but relatively large D’ values among variants in the minimum subsets (**Suppl. Fig. S5**).

Next, we evaluated the associations of the reduced rare lead haplotypes with BCa risk using the DRIVE data. Using the “Original haplotype calls” scheme (see Methods), two reduced rare lead haplotypes (Locus ID #5: 8 variants, length = 1.40 Mb, odds ratio (OR) = 1.50, Likelihood Ratio Test (LRT) *P* = 0.012; Locus ID #13: 3 variants, length = 98 kb, OR = 2.93, LRT *P* = 0.020) showed associations with BCa risk, adjusting for age and the first ten genotype PCs (**Table 3**). For the remaining 11 reduced rare lead haplotypes with LRT *P*>0.05, we explored two modified haplotype calls where risk-haplotype carriers were called using a subset of variants in the minimal set with the fewest imputation errors. Using the “High-confidence haplotype calls” scheme (see Methods), two additional reduced rare lead haplotypes of Locus ID #17 (7 variants, length = 201 kb, OR = 2.19, LRT *P* = 0.036) and Locus ID #18 (5 variants, length = 458 kb, OR = 7.35, LRT *P* = 0.019) showed BCa-risk associations. Furthermore, allowing one variant to have possibly been imputed poorly, the “High-confidence haplotype calls with one exception” scheme (see Methods), two additional reduced rare lead haplotypes on Locus ID #15 (4 variants, length = 318 kb, OR = 1.48, LRT *P* = 0.007) and Locus ID #16 (5 variants, length = 232 kb, OR = 1.54, LRT *P* = 0.030) showed associations with BCa risk (**Table 2**). By allowing a one-variant exception, the frequencies of these two lead haplotypes increased, resulting in attenuated but statistically significant effect sizes on BCa risk. Taken together, six of the 13 reduced rare lead haplotypes showed successful generalizability in the DRIVE imputed data.

**Table 3.** Generalizability Evaluation Results of 6 Reduced Lead Rare Haplotypes in the DRIVE Data with TOPMed Imputation.

| Locus | Lead Haplotype <sup>a</sup> | # Variants <sup>b</sup> | UK Biobank Combined Analysis |  |  |  | Generalizability Analysis Using the DRIVE Imputed Data |  |  |  |  |  |  |
| --- | --- | --- | --- | --- | --- | --- | --- | --- | --- | --- | --- | --- | --- |
|  |  |  | Frq0 <sup>c</sup> | Frq1 <sup>c</sup> | HR <sup>d</sup> | *P*<sub>Cox</sub> <sup>e</sup> | Exemption <sup>f</sup> | *C* <sup>g</sup> | Frq0 <sup>c</sup> | Frq1 <sup>c</sup> | OR <sup>d</sup> | *P*<sub>Fisher</sub> <sup>e</sup> | *P*<sub>LRT</sub> <sup>e</sup> |
| Analysis of the original haplotype calls in the Minimac4 output |  |  |  |  |  |  |  |  |  |  |  |  |  |
| 5 | 22_3362_3611 | 8 | 12.208 | 31.073 | 2.48 | 1.3×10<sup>-11</sup> | / | / | 11.273 | 17.130 | 1.505 | 0.013 | 0.0120 |
| 13 | 16_13434_13533 | 3 | 2.471 | 9.988 | 3.76 | 2.1×10<sup>-8</sup> | / | / | 0.989 | 3.160 | 2.939 | 0.014 | 0.0200 |
| Extended analysis I with high-confidence haplotype calls |  |  |  |  |  |  |  |  |  |  |  |  |  |
| 17 | 2_4129_4178 | 7 | 15.986 | 32.183 | 2.10 | 1.8×10<sup>-8</sup> | / | 0.020 | 1.780 | 3.991 | 2.191 | 0.036 | 0.0363 |
| 18 | 10_7217_7316 | 5 | 2.587 | 10.543 | 3.78 | 7.3×10<sup>-9</sup> | / | 0.002 | 0.198 | 1.330 | 7.353 | 0.045 | 0.0199 |
| Extended analysis II with high-confidence haplotype calls and 1-variant exemption |  |  |  |  |  |  |  |  |  |  |  |  |  |
| 15 | 13_4032_4131 | 4 | 2.994 | 11.098 | 3.57 | 1.3×10<sup>-8</sup> | 1 | 0.050 | 14.833 | 21.288 | 1.48 | 0.016 | 0.0077 |
| 16 | 4_27513_27612 | 5 | 2.645 | 9.433 | 3.26 | 1.2×10<sup>-6</sup> | 5 | 0.410 | 7.515 | 11.642 | 1.54 | 0.033 | 0.0300 |

<sup>a</sup> The lead haplotype is labeled as "Chromosome number\_index of the 1<sup>st</sup> variant\_index of the last variant".
<sup>b</sup> The number of imputed variants (Rs<sub>q</sub>>0.80 in the TOPMed imputation) among the variants in the haplotype window of size W after reduction using recursive partitioning (minsplit size=40; CP= $1 \times 10^{-10}$ ).
<sup>c</sup> Frq 0 and Frq1, haplotype frequencies (per 10,000) in breast cancer (BCa) controls and BCa cases of the respective dataset, respectively
<sup>d</sup> HR/OR, hazard ratio/odds ratio of BCa comparing women with vs. without the haplotype.
<sup>e</sup> $P_{\text{Cox}}/P_{\text{Fisher}}/P_{\text{LRT}}$ , p-value from Cox regression (with age taken as the time axis)/Fisher's exact test/likelihood-ratio test from logistic regression for testing the null hypothesis of no BCa-haplotype association (HR or OR=1.0), adjusting for ten PCs in the Cox and logistic regression.
<sup>f</sup> The index of the exempted variant on the reduced haplotype. /: not applicable or data not available
<sup>g</sup> $c$ , HDS error threshold used to define high-confidence haplotypes, subject to the constraint $\sum HDSE < c$ and $0 \leq c \leq 0.50$ .

## Discussion

Through systematically assessing BCa-haplotype associations genome-wide in one of the largest publicly available cohorts with GWAS SNP-array data, the UKBB, our analysis identified and replicated 13 rare haplotype associations with BCa risk, of which six BCa-haplotype associations were found generalizable in DRIVE case-control data. Note that the study is far from a comprehensive assessment of BCa-haplotype associations, especially because it used only ∼670,000 autosomal variants genotyped on the UKBB Axiom Array (see below for a more comprehensive discussion of the limitations of our study). Nevertheless, our results have critical implications on the germline autosomal genetic contributions to BCa risk.

The 13 replicated, rare risk haplotypes span multiple LD blocks (**Fig. 3)** and contain variants whose minor alleles are relatively common. But the combination of alleles across the LD blocks, i.e., the haplotype, is rare. LD analysis showed consistently low *r*^2^ but relatively high D’ values among the alleles of these haplotypes in both UKBB and DRIVE datasets (**Suppl. Fig. S5**). These observations indicate that it is not the individual common alleles (or their chromosomal segments) in regions with occasional recombination (high D’ values), but their specific combinations occurring infrequently (low frequency and *r*^2^), that are associated with the large elevation of BCa risk. Consistent with this finding, the BCa-risk-associated haplotypes identified using smaller windows were common, located in a single LD block, and often tagged by well-known, common, single risk variants. Our analysis demonstrates the association of this specific configuration of common germline variants with BCa risk, as well as the feasibility of utilizing existing GWAS SNP-array datasets in assessing haplotype-disease associations for identifying such rare risk haplotypes.

To support the notion that specific combinations of common alleles (or their segments) are risk elevating, the functional annotations showed many of the variants on the 13 replicated haplotypes have indications for regulatory roles for gene expression, enhancer activity, DNA-motif change, and 3D chromatin interactions. The specific rare combinations of alleles may represent “unfortunate” recombination results with regulatory consequences influencing BCa risk. For example, the haplotype on 22q12.1 (Locus #5, **Fig. 2A**), one of the two rare risk haplotypes that were found generalizable in DRIVE and mapped to known BCa-risk genes (the other was Locus # 15 discussed below), spans a 1.4 Mb region with dozens of genes, the majority of which have variants reported by GWAS for their associations with BCa risk or BCa-specific mortality (3,13), including *MN1* encoding a transcription regulator and an oncogene (14) and *CHEK2* involving in the DNA damage repair and cell cycle by interacting with BRCA1/2 and other proteins *(15)*. However, it remains elusive whether these GWAS risk variants influence BCa risk through common or independent mechanisms. Our exploration of potential regulatory roles for the 250 variants on the haplotype found (**Suppl. Table S6**): 1) 12 are cis-eQTLs of four genes (*TTC28, TTC28-AS1, KREMEN1*, and *CTA-292E10*.*6*/*lnc-CCDC117-2*) in the GTEx breast mammary tissue (16); 2) 34 are enhancer variants and possibly regulate ten genes (*MN1, TTC28, CHEK2, ZNRF3, XBP1, HSCB, CCDC117, EWSR1, RHBDD3*, and *KREMEN1)* via enhancer-promoter interactions in BCa-related cells (17,18); 3) 15 are predicted to affect the binding of 17 unique DNA motifs (19); 4) 69 reside in the anchor regions of chromatin loops or TADs boundaries, reflecting their role in regulating local chromatin structures (20,21). The rare haplotype on 13q13.1 (Locus #15) mapped to a 361 kb region with multiple genes such as *FRY, ZAR1L*, and *BRCA2* (**Fig. 2H**). Besides, five genes (*N4BP2L1/2, PDS5B, RP11-37E23*.*5*, and *RP1-257C22*.*2*) downstream of *BRCA2* were also predicted to be targets of eQTLs or enhancer variants on the haplotype in Locus #15. *FRY* encodes a microtubule-binding protein that is essential to mammary gland development and affects BCa cell functions (22). *ZAR1L* encodes an RNA regulator that can inhibit *BRCA2* transcription in a cell cycle-dependent manner (23). APRIN (PDS5 cohesin-associated factor B) encoded by *PDS5B* is a regulator of the cohesin complex and can interact with BRCA2 in the DNA damage repair and carcinogenesis (24). In summary, the functional annotations showed the following shared features across the 13 replicated, rare risk haplotypes: 1) most regions overlap TAD boundaries and are enriched in chromatin loops; 2) most of their variants are predicted to alter motif binding (17-83 unique DNA motifs with stronger binding and 8-73 unique motifs with weaker binding); and 3) variants located near their 5’ or 3’ ends are more critical to the observed risk effects (**Suppl. Fig. S2**). These features support our speculation that the rare risk haplotypes we identified represent long-range interactions with regulatory consequences influencing BCa risk.

Alternatively, the rare combination of common alleles may constitute a haplotype on which a risk-altering mutation arose, and the haplotypes we identified may be approximate tags of such mutations. For example, upon searching the TOPMed-imputed data generated on 181,034 UKBB women, we found that all 108 (100%) carriers of the Locus #15 rare haplotype carry a BRCA2 truncating mutation p.K3326X, which, in contrast, is carried by only 3,201 (1.7%) of 180,926 non-carriers of the haplotype (Fisher’s exact *P*=3.4×10^−189^): the corresponding frequencies in the DRIVE TOPMed-imputed data were 63.0% of the haplotype carriers vs. 1.7% of the non-carriers of the haplotype (Fisher’s exact P=1.3×10^−45^). Thus, both UKBB and DRIVE data show strong correlation between the Locus #15 haplotype and the p.K3326X mutation. Note that this specific BRCA2 mutation has been indicated as a mutation associated with increased BCa risk (25), but its role in BCa has not been functionally characterized (26). We hypothesize that the rare risk haplotype of Locus #15 captures the risk-increasing effect of BRCA2 p.K3326X itself, which might be only seen in the presence of other mutations in the region. Further investigation with deep sequencing of mutational changes in rare haplotypes carriers would be able to distinguish the two possible causal factors the risk haplotypes represent.

The number of risk haplotypes we found was small despite our genome-wide analysis. To interpret this result, we estimated statistical power of the entire three-stage analysis (discovery/replication with the UKBB data and generalizability evaluation with the DRIVE data). The approximate power for each of the six generalizable, rare risk haplotypes was in the range of 1.3-30.6% (**Suppl. Table S7**). These power estimates suggest that there are additional 2-78 generalizable, rare risk haplotypes we did not detect with similar frequencies and risk effects as those that passed all three stages of our analysis. In addition, the UKBB Axiom Array contained a relatively small number of variants. Thus, the haplotypes we examined are a small subset of all haplotypes that can be considered by WGS. Extrapolation of our results suggests that WGS data on sufficiently large size cohorts and case-control studies should lead to the discovery of additional rare risk haplotypes as well as rare risk variants.

Aside from the two risk haplotypes (Loci #5 and #15) above, two other generalizable, rare risk haplotypes have evidence for relevance to BCa risk. The haplotype on 4q28.3 contained a few long noncoding RNAs but no protein-coding gene (Locus #16, **Fig. 2I**). Its lead variant rs116648919 was in LD with downstream variant rs6837069 (*r*^2^<0.01 and D’=1.0, **Table 2**) which was associated with BCa-specific mortality (OR=1.92, *P*=1×10^−6^) in a 2019 GWAS (11). The haplotype on 10p12 falls into the region of *KIAA1217* (Locus #18, **Fig. 2K**). Its lead variantrs10764446, located in intron 2 of *KIAA1217*, is in modest LD with another *KIAA1217* intronic variant rs11013833 (*r*^2^=0.53 and D’=0.85) which was reported to increase the risk of the breast-colorectal cancer phenotype (OR=1.37, *P*=6×10^−6^) in a 2018 GWAS (10). *KIAA1217* belongs to the human large KIAA gene family and its expression is highest in female breast mammary tissue among many cell types stratified by sex, according to the GTEx RNA-seq data (16). RNA-seq analysis conducted by Asmman et al. (27) detected a translocation t(10;14)(p12;q32) in estrogen receptor (ER)-positive BCa tumor, causing the full coding sequence and 3’TUR of *SERPINA1* were fused into the 5’-end of *KIAA1217*. As such, the fusion gene would not produce KIAA1217 protein and might alter the transcription stability of *SERPINA1*. A 2015 study by Chan et al. identified an ER binding site in *SERPINA1* promoter and *SERPINA1* transcription was estradiol dependent. These data imply that the rare haplotype of Locus #18 may contribute to BCa risk by affecting the transcription outcomes of *KIAA1217*. Although these two loci were implicated for BCa risk previously, the frequency and effect sizes we observed here for their risk haplotypes (HR estimates exceeding 3.4 in our replication analysis with Locus #18’s OR estimate exceeding 7.3 in DRIVE) support the rare-variant/haplotype model over the common-variant counterpart. We could not find any previous reports linking BCa and the remaining two generalizable, rare risk haplotypes, one on 16q21 (Locus #13, **Fig. 2F**) and the other on 2p24.3 (Locus #17, **Fig. 2J**).

Our haplotype analysis relied on phasing accuracy. While both UKBB and DRIVE are large studies, phasing/imputation uncertainty in calling haplotypes, especially rare haplotypes, is a major concern. However, phasing/imputation errors should affect both BCa cases and BCa-free women indiscriminately, lowering the power of all three stages of our analysis. While this issue might explain why seven (Locus IDs: # 6, 9, 11, 12, 14, 19, and 20) of the 13 replicated, rare haplotypes were not found generalizable in DRIVE, it should not negate the validity of the six found to be generalizable. The above problem can be partially addressed by incorporating phase information from WGS reads into haplotype estimation or by using advanced sequencing technologies such as long-read or single-cell sequencing (28,29). Our crude approach to this problem was to use well-imputed variants only, possibly combining with exempting one variant from a haplotype membership in the DRIVE analysis, to account for potential errors in its phase or dosage. By exempting one variant, rare haplotypes were called more liberally with increased frequencies that did not precisely match the original haplotypes’, contributing to attenuated OR estimates.

Our analysis was restricted to women of “white British” in the discovery and replication, and women of broader European genetic ancestry in the generalizability evaluation. These were a methodological necessity because populations of different ancestries differ in allele frequencies and LD structures, which could confound the BCa-haplotype associations. No population of non-European ancestries has a large BCa GWAS dataset of the size that is comparable to UKBB and DRIVE. This disparity is an important challenge our field must tackle. Also, we did not consider BCa subtypes (e.g., hormone receptor subtypes): risk haplotypes specific to a particular BCa subtype (2) may exist.

In conclusion, we have leveraged a large “white British” GWAS cohort data to discover and replicate 13 long, rare haplotypes associated with BCa risk through a genome-wide two-stage haplotype analysis, of which six were found generalizable in an independent case-control study of women with European genetic ancestry. Our findings highlight the contribution of rare haplotypes to BCa risk and suggest that the rare risk effects are possibly of a regulatory nature characterized by long-range interactions. We expect that the ability to assess rare variant or haplotype associations will be greatly improved when large-scale sequencing data become available. Further efforts are required to characterize the exact causal mechanisms of the rare risk haplotypes. Finally, the existence of rare BCa-risk haplotypes is consistent with the patterns of BCa incidence observed in large-scale epidemiological twin and family studies (Yasui et al. submitted as a separate manuscript) and supports the genetic-heterogeneity/rare-variant hypothesis of BCa (6), which attributes an individual’s disease risk elevation to one of many rare, risk variants/haplotypes.

## Data Availability

The data generated in this study are available within the article and its supplemental data files. Individual-level genotype and phenotype data are publicly available by submitting request to the UK Biobank and dbGaP (study accession: phs001265.v1.p1).

## Acknowledgments

We thank the high-performance computing facility at St. Jude Children’s Research Hospital for computational support. This study has been conducted using the UK Biobank Resource under Application Number 44891. OncoArray genotyping and phenotype data harmonization for the Discovery, Biology, and Risk of Inherited Variants in Breast Cancer (DRIVE) breast-cancer case control samples was supported by X01 HG007491 and U19 CA148065 and by Cancer Research UK (C1287/A16563).

## Financial support

This work is supported by National Cancer Institute Grant R01 CA216354 (W.M., W.L., Y.S., C.L., J.L.B., L.R., and Y.Y.), American Lebanese Syrian Associated Charities (F.W., W.M., W.L., Y.S., Z.W., J.L.B., L.R., and Y.Y.), and Alberta Machine Intelligence Institute (C.L. and Y.Y.).

## Declaration of conflict interests

The authors declare no potential conflicts of interest.

## Notes

### Competing Interest Statement

The authors have declared no competing interest.

### Funding Statement

National Cancer Institute Grant R01 CA216354, American Lebanese Syrian Associated Charities, and Alberta Machine Intelligence Institute.

### Author Declarations

1. UK Biobank: https://www.ukbiobank.ac.uk/ 2. DRIVE study: https://www.ncbi.nlm.nih.gov/projects/gap/cgi-bin/study.cgi?study_id=phs001265.v1.p1

