## Supplementary Data for "Genome-wide Analysis of Rare Haplotypes Associated with Breast Cancer Risk: Discovery, Replication, and Generalizability Evaluation"

**The supplementary data includes seven tables and five figures.**

Table S1. Permutation Experiment Estimating the False Discovery Rate of the Two-Stage Haplotype Analysis Using the UKBB Phased Data 2

Table S2. List of 436 Haplotypes Conferring BCa Risk Identified by the Two-Stage Analysis of the UKBB Phased Data 2

Table S3. Seven Common Haplotype Risk Loci Identified in the UKBB Phased Data 3

Table S4. Investigation of Critical Regions Of the 13 Rare Risk Haplotypes Detected in the UKBB Phased Data 4

Table S5. Functional Annotation of SNPs on the 13 Rare Risk Haplotypes Detected in the UKBB Phased Data 4

Table S6. Haplotype-level Summary of Variants Exhibiting Putative Regulatory Roles 4

Table S7. Statistical Power of Six Rare Risk Haplotypes Detected in The Two-stage Analysis Plus the Generalizability Evaluation 5

Figure S1. Genetic Ancestry of 181,034 White British Women Included in the Discovery-Replication Analysis 6

Figure S2. Investigation of Critical Regions of 13 BCa Rare Haplotypes Identified in the UKBB Phased Data 7

Figure S3. Sample-level Quality Control of 60,015 Women in the DRIVE study 8

Figure S4. Quality of Imputed Variants on the 13 Rare Haplotypes in the DRIVE TOPMed Imputed Data 9

Figure S5. LD Pattern of 13 Reduced Rare Haplotypes Selected for Generalizability Evaluation 10

**Table S1. Permutation Experiment Estimating the False Discovery Rate of the Two-Stage Haplotype Analysis Using the UKBB Phased Data**

| Window Size [a] | Permuted Dataset [b] | No. of Windows | Genomic Length | Retrospective Discovery | | Prospective Replication | |
|---|---|---|---|---|---|---|---|
| | | | | Total Tests [c] | $P<5\times10^{-8}$ | $P<0.01$ | No. of Loci [d] |
| W=5 | 1 | 646,336 | 17.2 Kb | 4,803,745 | 3 | 0 | 0 |
| | 2 | 646,325 | 17.2 Kb | 4,803,094 | 0 | 0 | 0 |
| | 3 | 646,321 | 17.2 Kb | 4,801,281 | 3 | 0 | 0 |
| W=10 | 1 | 646,248 | 38.8 Kb | 12,240,197 | 18 | 0 | 0 |
| | 2 | 646,248 | 38.8 Kb | 12,239,174 | 26 | 0 | 0 |
| | 3 | 646,247 | 38.8 Kb | 12,236,774 | 34 | 0 | 0 |
| W=20 | 1 | 646,028 | 81.8 Kb | 34,466,125 | 117 | 0 | 0 |
| | 2 | 646,028 | 81.8 Kb | 34,479,353 | 243 | 0 | 0 |
| | 3 | 646,028 | 81.8 Kb | 34,469,414 | 196 | 0 | 0 |
| W=30 | 1 | 645,808 | 124.9 Kb | 58,118,695 | 348 | 0 | 0 |
| | 2 | 645,808 | 124.9 Kb | 58,122,202 | 545 | 2 | 1 |
| | 3 | 645,808 | 124.9 Kb | 58,142,781 | 459 | 5 | 1 |
| W=50 | 1 | 645,368 | 211.0 Kb | 88,444,047 | 902 | 9 | 4 |
| | 2 | 645,368 | 211.0 Kb | 88,408,290 | 937 | 4 | 2 |
| | 3 | 645,368 | 211.0 Kb | 88,510,134 | 1,135 | 8 | 1 |
| W=100 | 1 | 644,229 | 426.4 Kb | 85,914,872 | 1,191 | 1 | 1 |
| | 2 | 644,216 | 426.4 Kb | 85,846,558 | 1,753 | 7 | 1 |
| | 3 | 644,265 | 426.4 Kb | 86,075,748 | 1,856 | 1 | 1 |
| W=250 | 1 | 616,766 | 1.08 Mb | 25,851,440 | 1,018 | 0 | 0 |
| | 2 | 616,809 | 1.08 Mb | 25,818,502 | 1,196 | 0 | 0 |
| | 3 | 617,309 | 1.08 Mb | 25,969,778 | 1,513 | 0 | 0 |

[a] The number of consecutive genotyped variants that define the haplotypes. Each window is shifted by one variant.

[b] The BCa status and other covariates (i.e., age of onset, age at enrollment, and top ten genotype principal components) of each woman were concatenated as a vector during each permutation.

[c] The total number of haplotypes with frequency greater than 0.1% among 5,487 pre-enrollment BCa cases.

[d] Independent loci were determined by the PLINK LD clumping with $r^2>0.10$ and a maximum distance of 500 kb between the lead haplotype and the other correlated ones.

**Table S2. List of 436 Haplotypes Conferring BCa Risk Identified by the Two-Stage Analysis of the UKBB Phased Data**

This is a long table. Pease refers to an external Excel file “**Suppl Tables.xlsx**.”

**Table S3. Seven Common Haplotype Risk Loci Identified in the UKBB Phased Data**

| Haplotype Window | | | | UKBB Combined Analysis | | | | GWAS with All UKBB Women | | | | | Nearest Genes | GWAS Catalog [h] |
|---|---|---|---|---|---|---|---|---|---|---|---|---|---|---|
| Locus | Hits [a] | Lead Haplotype [b] | W [c] | Frq0 [d] | Frq1 [d] | HR [e] | $P_{cox}$ [f] | Variant | EA [g] | EAF [g] | HR [e] | $P_{cox}$ [f] | | |
| 1 | 80 | 10_27798_27802 | 5 | 0.369 | 0.433 | 1.30 | $2.3\times10^{-67}$ | rs2981575 | A | 0.606 | 0.77 | $4.1\times10^{-66}$ | *FGFR2* | Previously reported: OR=1.29, *P*=$2.3\times10^{-54;}$ PubMed: 32808324 |
| 2 | 84 | 16_11154_11158 | 5 | 0.229 | 0.271 | 1.25 | $2.1\times10^{-39}$ | rs4784227 | T | 0.239 | 1.25 | $5.6\times10^{-42}$ | *CASC16* | Previously reported: OR=1.23, *P*=$7.0\times10^{-201}$; PubMed: 29059683 |
| 3 | 16 | 2_44300_44304 | 5 | 0.488 | 0.521 | 1.14 | $3.8\times10^{-18}$ | rs13387042 | G | 0.500 | 0.88 | $3.1\times10^{-18}$ | *LINC01921, DIRC3-AS1* | Previously reported: OR=1.12, *P*=$1.0\times10^{-95;}$ PubMed: 23535729 |
| 4 | 39 | 11_16541_16545 | 5 | 0.106 | 0.128 | 1.24 | $1.2\times10^{-21}$ | rs559664 | G | 0.328 | 1.07 | $2.6\times10^{-05}$ | *LINC01488, CCND1* | In LD ($r^2$=0.158, D'=1) with rs78540526: OR=1.32, *P*=$1.0\times10^{-131}$; PubMed: 29059683 |
| 7 | 5 | 5_11672_11676 | 5 | 0.842 | 0.821 | 0.87 | $3.9\times10^{-13}$ | rs76485124 | T | 0.053 | 1.18 | $4.2\times10^{-08}$ | *C5orf67, MAP3K1* | In LD ($r^2$=0.234, D'=1) with rs62355901: OR=1.19, *P*=$3.0\times10^{-98}$; PubMed: 29059683 |
| 8 | 7 | 3_7827_7831 | 5 | 0.459 | 0.433 | 0.90 | $2.2\times10^{-12}$ | rs3920005 | C | 0.472 | 0.90 | $5.9\times10^{-12}$ | *NEK10* | In LD ($r^2$=0.984, D'=1.0) with rs60936670: OR=1.11, *P*=$8.0\times10^{-64}$; PubMed: 29059683 |
| 10 | 12 | 8_28003_28007 | 5 | 0.415 | 0.389 | 0.90 | $4.3\times10^{-12}$ | rs12541832 | A | 0.312 | 1.11 | $1.2\times10^{-11}$ | *CASC8, CASC21* | In LD ($r^2$=0.323 D'=0.924) with rs10096351: OR=1.11, *P*=2.0×10-64; PubMed: 32887889 |

[a] The number of correlated haplotypes at the same risk locus/region.

[b] The lead haplotype labeled as "Chromosome number_index of the 1st variant_index of the last variant, was defined as the one which showed the minimal Cox-regression p-value in the UKBB combined analysis.

[c] W denotes the length of haplotype, i.e., the number of consecutive genotyped variants.

[d] Frq 0 and Frq1 represent haplotype frequency in BCa-free women and BCa cases, respectively.

[e] HR: hazard ratio.

[f] $P_{cox}$, p-value from Cox regression for testing the null hypothesis no BCa-haplotype association (HR=1.0), with age taken as the time axis and adjusting for ten genotype PCs.

[g] EA: effect (or alternate) allele. EAF: effect allele frequency

[h] For each haplotype locus, we searched for known GWAS variants reported in the GWAS catalog within the 500-kb region of the lead variant.

**Table S4. Investigation of Critical Regions Of the 13 Rare Risk Haplotypes Detected in the UKBB Phased Data**
This is a long table. Please refer to an external Excel file "**Suppl Tables.xlsx.**"

**Table S5. Functional Annotation of SNPs on the 13 Rare Risk Haplotypes Detected in the UKBB Phased Data**
This is a long table. Please refer to an external Excel file "**Suppl Tables.xlsx.**"

**Table S6. Haplotype-level Summary of Variants Exhibiting Putative Regulatory Roles**

| Locus [a] | W [b] | Number of Annotated Variants [c] | | | | | Summary of Motif Changes at the Haplotype-level | | | Putative Targets [d] |
|---|---|---|---|---|---|---|---|---|---|---|
| | | eQTLs | Enhancers | TAD20K | Loops | Motifs | Unique changes | Increased binding (Top 3 motifs) | Decreased binding (Top 3 motifs) | |
| 5 | 250 | 18 (12) | 34 | 5 | 68 | 15 | 23 | 17 (NF-kappaB, BATF, ATF3) | 8 (ATF3, Smad3, E2F) | MN*, PITPNB, TTC28*,$^{+}$, KREMEN1 *,$^{+}$, TTC28-AS1 $^{+}$, CCDC117 *, CTA-292E10.8$^{+}$, CHEK2 *, CTA-292E10.6/lnc-CCDC117-2 *, HSCB *, XBP1 *, ZNRF3 *, EWSR1 *, RHBDD3 * |
| 13 | 100 | 5 (0) | 3 | 0 | 0 | 68 | 96 | 61 (Znf143, TLX1::NFIC, AP-4) | 50 (STAT, NF-kappaB, Pou2f2) | RP11-354I13.1, RP11-354I13.2, RP11-430C1.1 |
| 17 | 50 | 1 (0) | 4 | 0 | 0 | 34 | 66 | 38 (RXR::LXR, Irf, Ets) | 36 (Mef2, CDP, TAL1) | AC092635.1, LINC00276 |
| 18 | 100 | 17 (0) | 11 | 9 | 27 | 65 | 106 | 64 (Pou2f2, Irf, Bach1) | 65 (Foxc1, Pou2f2, ELF1) | KIAA1217 |
| 15 | 100 | 21 (1) | 5 | 3 | 16 | 70 | 109 | 65 (SP1, BAF155, Foxj2) | 70 (AIRE, Bach1, AP-1) | FRY, FRY-AS1, ZAR1L, BRCA2 *, N4BP2L1 *, N4BP2L2 *, PDS5B *, RP11-37E23.5 *, RP1-257C22.2$^{+}$, RP11-207N4.3, |
| 16 | 100 | 2 (0) | 0 | 3 | 0 | 74 | 126 | 83 (DMRT1, Gfi1b, Hand1) | 73 (Foxc1, RXR::LXR, Irf) | RP11-553P9.2, AC108867.1, RP11-192C21.2, RP11-780O17.1, RP11-553P9.3 |

[a] Locus, the index of rare haplotype locus.
[b] W, window size.
[c] eQTLs, the number of variants that are reported to be cis-eQTLs in any GTEx tissues (and breast mammary tissue). Enhancer, the number of variants located within active human enhancers. TAD, the number of variants falling within 20 kB of the boundary of a human topologically associated domain (TAD). Loop, the number of variants residing in the anchor regions of chromatin loops. Motifs, the number of variants predicted to alter motif binding with LOD score of difference between two alleles is above 3 (see Methods for details).
[d] Putative targets were identified based on the evidence of eQTLs (marked by *), enhancer-promoter interactions (marked by $^{+}$), and genomic mapping (nearest genes).

**Table S7. Statistical Power of Six Rare Risk Haplotypes Detected in The Two-stage Analysis Plus the Generalizability Evaluation**

| Locus [a] | Lead Haplotype [b] | W [c] | Frq0 [d] | Frq1 [d] | Effect Size [e] | Power [f] | Expected No. of Causal Haplotypes |
|---|---|---|---|---|---|---|---|
| 5 | 22_3362_3611 | 250 | 6.046 | 22.750 | 3.43 | 0.306 | 3 |
| 13 | 16_13434_13533 | 100 | 1.918 | 9.433 | 3.77 | 0.023 | 43 |
| 17 | 2_4129_4178 | 50 | 9.505 | 23.305 | 2.08 | 0.013 | 79 |
| 18 | 10_7217_7316 | 100 | 2.035 | 9.988 | 3.41 | 0.017 | 58 |
| 15 | 13_4032_4131 | 100 | 2.529 | 11.098 | 3.34 | 0.023 | 44 |
| 16 | 4_27513_276120 | 100 | 1.889 | 9.433 | 3.55 | 0.017 | 58 |

[a] Locus, the index of rare haplotype locus.
[b] Lead haplotype, labeled as "Chromosome number_index of the 1st variant_index of the last variant" and defined as the haplotype which showed the minimal Cox p-value in the UKBB combined analysis.
[c] W, haplotype window size.
[d] Frq0 and Frq1, haplotype frequencies among 172,023 BCa-free women and 9,011 BCa cases in the UKBB dataset.
[e] Unbiased haplotype risk effect size estimated by the UKBB prospective replication analysis.
[f] Approximate power for detecting a rare haplotype of the same frequency, effect size, and statistical significance in all three phases of the study: discovery (5,487 cases vs. 175,547 controls with $p < 5\times 10^{-8}$), replication (3,524 cases vs. 172,023 controls with $p < 0.01$), and generalizability evaluation (30,064 cases vs. 25,282 controls with $p < 0.05$). The power analysis was performed based on Fisher's Exact Test using the R package 'statmod.'

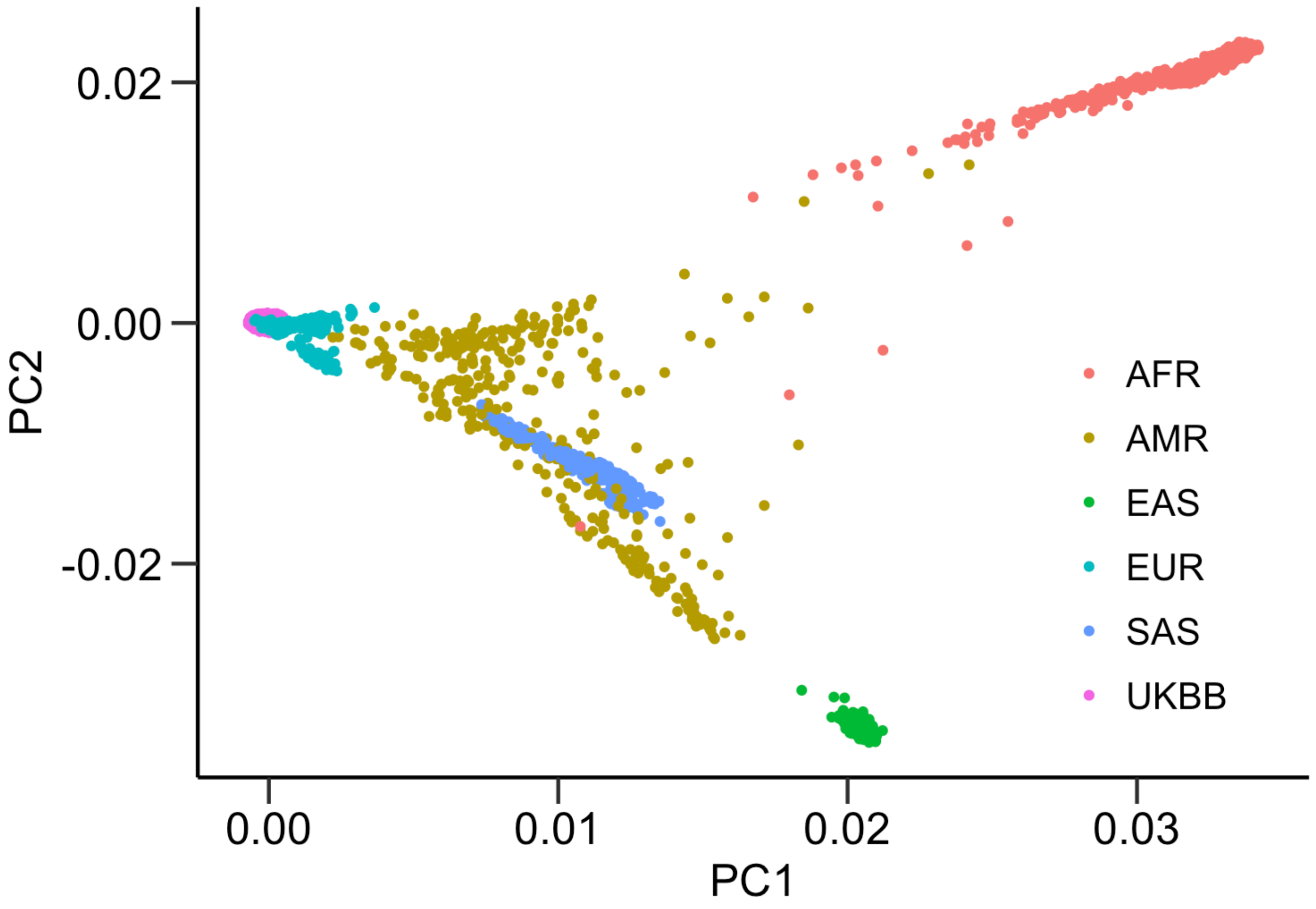


**Figure S1. Genetic Ancestry of 181,034 White British Women Included in the Discovery-Replication Analysis**
The genetic background of 181,034 White British women selected from the UKBB source was evaluated by principal components analysis (PCA) using a set of 283,166 LD-pruned variants ($r^2$>0.5 and a window size of 50 SNPs). The genomic similarity between UKBB and 1000Genomes subjects is visualized by plotting the first principal component (PC1) against PC2. AFR, African. AMR, Ad Mixed American. EAS, East Asians. SAS, South Asians. EUR, European.

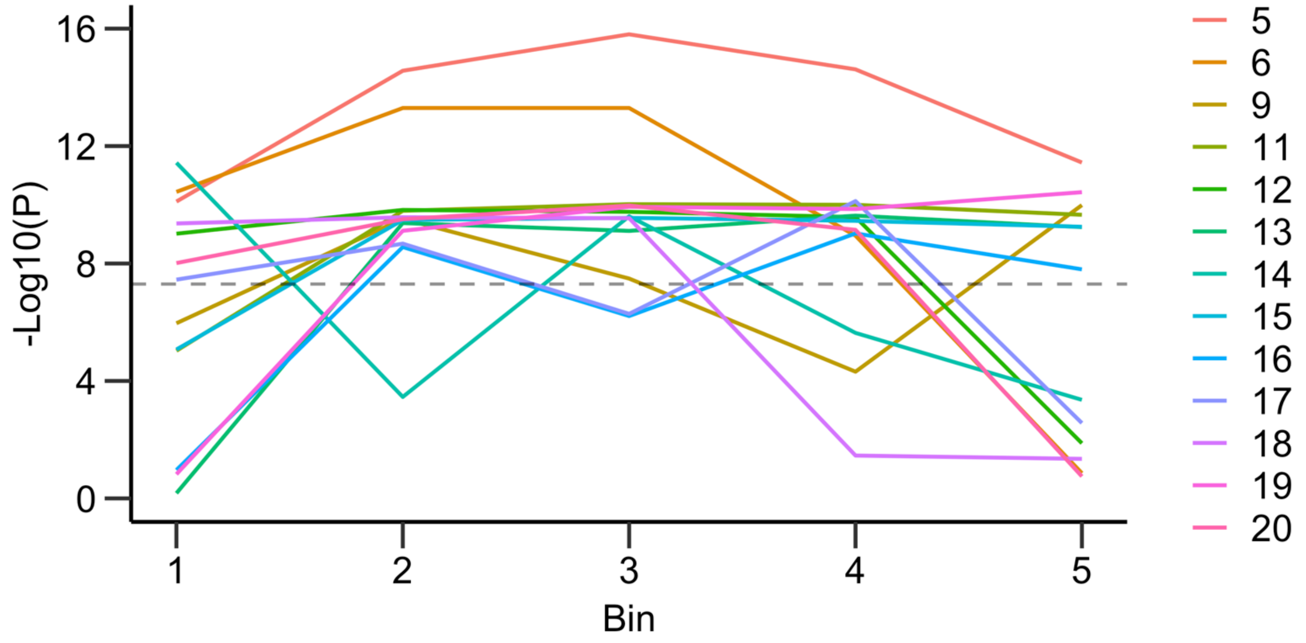


**Figure S2. Investigation of Critical Regions of 13 BCa Rare Haplotypes Identified in the UKBB Phased Data**
SNPs on each haplotype were split into five bins, each with an equal number of consecutive variants. For example, Bin=1 and 5 are the first and last 20% of variants, respectively. The genetic association between rare haplotypes and BCa risk was evaluated by excluding each bin sequentially. Colored lines represent 13 rare loci; the indexes shown in the legend are consistent with those in Table 1.

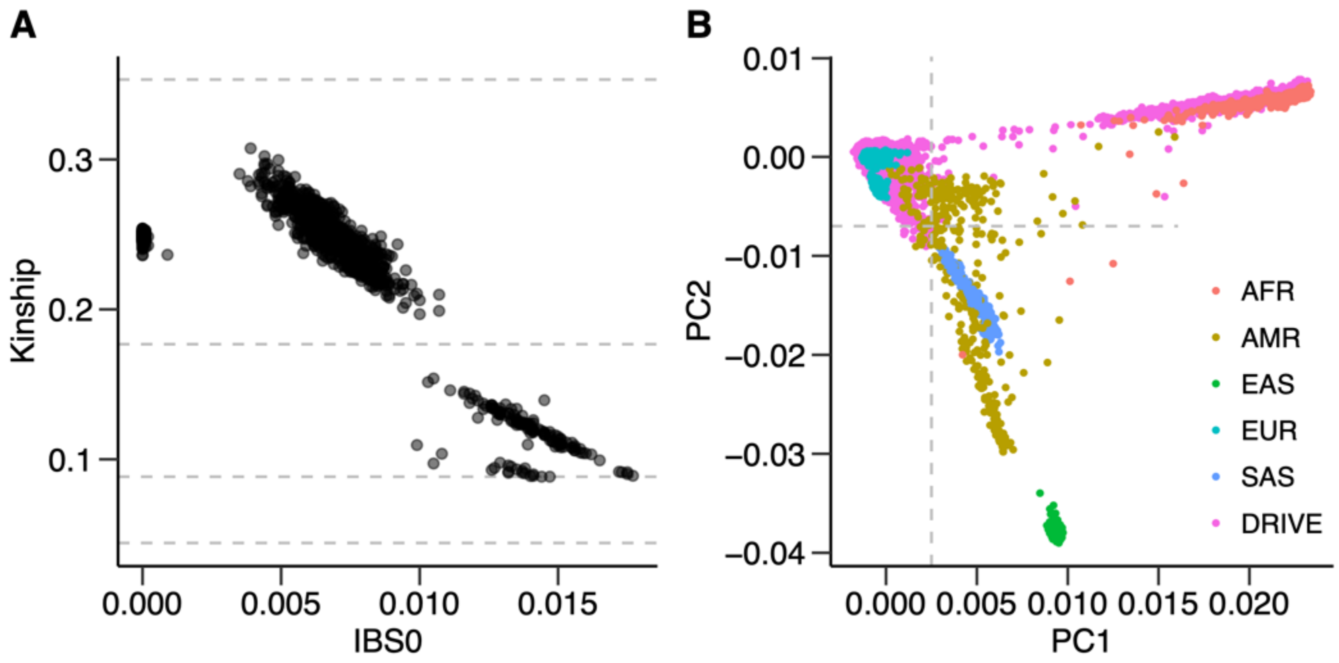


**Figure S3. Sample-level Quality Control of 60,015 Women in the DRIVE study**
(A) Scatter plot of kinship coefficients and IBS0 statistics among 1,696 pairs who were identified to be relatives, i.e., kinship coefficient >0.084. (B) The genetic ancestry of 59,958 DRIVE women was evaluated by PCA. PC1 and PC2 denote the first two principal components. Note that 57 women with high X chromosome homozygosity estimates (i.e., F-statistic>0.3) were excluded before PCA. A subset of 55,450 women of European ancestry were identified based on PC1 ≤0.0025 and PC2 ≥-0.007, as indicated by vertical and horizontal dotted lines. The 1000 Genomes Phase 3 data consists of five major populations: African (AFR), Ad Mixed American (AFR), East Asians (EA), South Asians (SAS), and European (EUR).

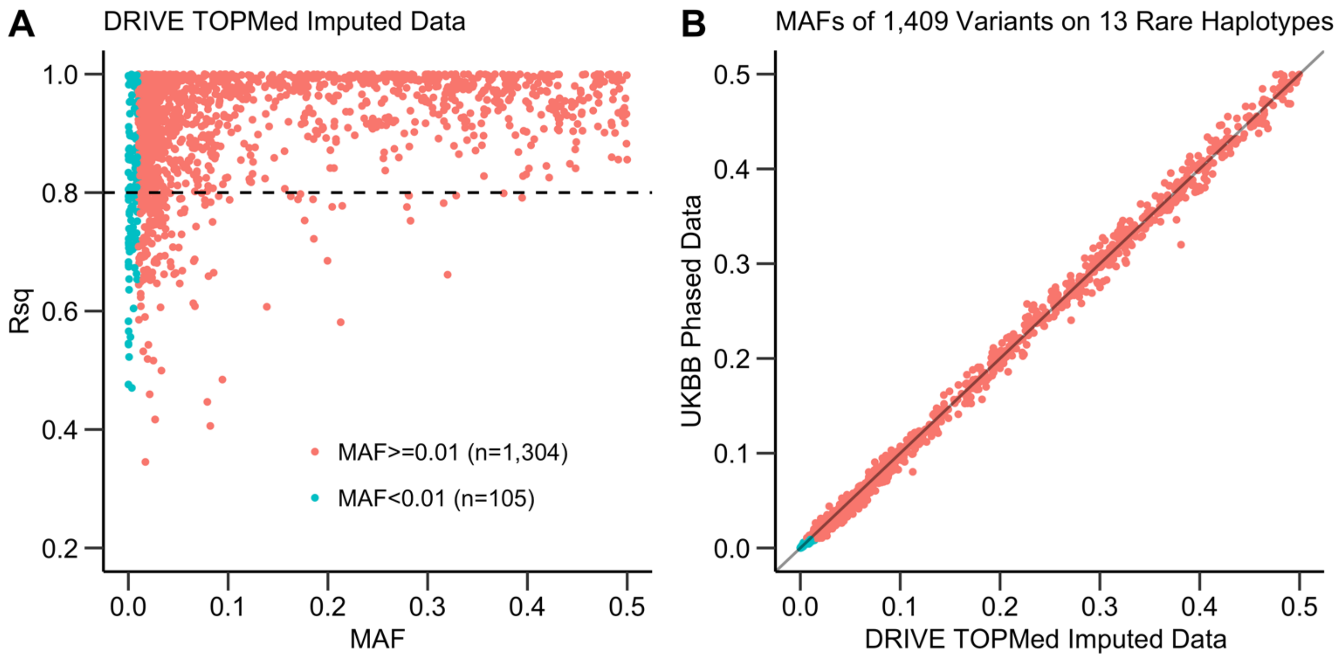


**Figure S4. Quality of Imputed Variants on the 13 Rare Haplotypes in the DRIVE TOPMed Imputed Data**
(A) The scatter plot shows the Minimac4 imputation scores (Rsq) and minor allele frequencies (MAFs) of 1,409 variants that were initially filtered based on Rsq ≥0.30 among women in the DRIVE study. Of these variants, 1,304 met Rsq≥0.80 as indicated by the horizontal dashed line. (B) The scatter plot shows MAFs of 1,409 variants in the UKBB phased data (y-axis) versus the DRIVE imputed datasets (x-axis).

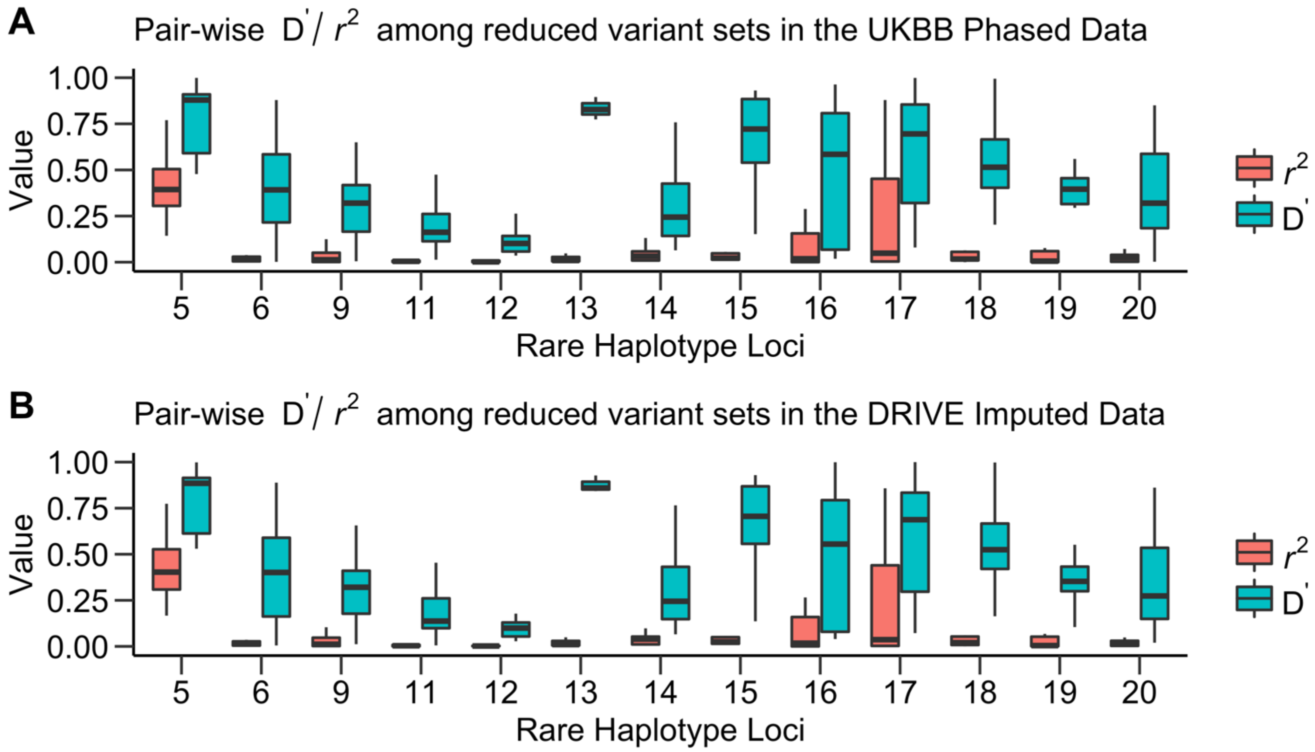


**Figure S5. LD Pattern of 13 Reduced Rare Haplotypes Selected for Generalizability Evaluation**
(A) For each rare haplotype, pair-wise D' and $r^2$ values (y-axis) for the reduced variant sets were computed among 181,034 White British women from the UKBB cohort. The indexes of 13 rare haplotype loci (x-axis) are consistent with those in Table 1. (B) Pair-wise *D'* and $r^2$ values for the same reduced variant sets were computed among 55,346 women of European ancestry from the DRIVE study.
